# A Systematic Review of Psychological Resilience in the COVID-19 Responses: Current Research and Future Directions

**DOI:** 10.1101/2025.02.15.25322337

**Authors:** Atena Pasha, Abdul-Hanan Saani Inusah, Jannatun Nayem, Xiaoming Li, Shan Qiao

## Abstract

**Highlights:**

- The COVID-19 pandemic has highlighted the importance of psychological resilience for well-being
- Psychological resilience was associated with multi-level socio-ecological factors.
- CD-RISC was the most used tool for measuring resilience during COVID-19.
- mHealth, social support, and policy reform can improve future public health crisis preparedness.

**Background:** The COVID-19 pandemic has highlighted the importance of psychological resilience for well-being. However, no systematic review has been synthesizing global literature on this topic. This review examines psychological resilience across diverse populations, focusing on measurement, associated factors, and future public health preparedness strategies.

**Methods:** Guided by the PRISMA, a thorough search of PsycINFO, PubMed, ScienceDirect, and Web of Science was conducted in February 2024, including quantitative studies considering psychological resilience during the COVID-19 pandemic published in peer-reviewed English journals from 2020 to 2024, using search terms relevant to psychological resilience and COVID-19.

**Results:** Sixty-eight studies were included, mostly using CD-RISC to measure psychological resilience. Multiple factors at various socio-ecological levels were associated with psychological resilience. During the pandemic, effective strategies in promoting resilience included promoting social connectedness through government policies and health communication campaigns and implementing personalized interventions such as telehealth services. For future public health crises preparedness, long-term strategies emphasized investments in mental health services, resilience-building activities, and providing robust support for mental health across diverse populations. Community and educational initiatives should focus on mental health promotion programs in colleges and schools and life skills training to improve interpersonal skills and job crafting. Technological solutions like remote consultations, mHealth applications, and telehealth were also recommended.

**Conclusion:** This review suggests that future public health crises preparedness requires prioritizing investments in mental health services, resilience-building activities, and mHealth applications. These insights provide a foundational resource for research, policy, and interventions to strengthen resilience globally at individual, community, and structural levels.

## 1. INTRODUCTION

The global public health crisis caused by COVID-19 has imposed a significant psychological impact on individuals ^1–3^. The resulting social isolation and economic uncertainty have led to a marked increase in mental health concerns, including feelings of loneliness, anxiety, depression, and suicidal thoughts, almost for all age groups ^4^. Older adults are at increased risk for the toxic effects of the virus and may have less access to virtual communication ^5^. Young people, who are already at a higher risk of mental health problems due to their age, have shown elevated levels of mental health challenges ^6, 7^. College students, who generally experience higher rates of mental health issues and lower well-being, have displayed increased mental health struggles since the start of the pandemic ^8–10^. Furthermore, the pandemic has posed significant challenges for mental health services to operate effectively ^1–3, 11, 12^. Therefore, researchers and policymakers predicted a significant rise in mental health issues due to the extensive measures implemented ^13^.

However, people exhibit diverse responses to the challenges they face ^14^. Extensive research has provided compelling evidence on how resilience may help individuals positively cope with major negative life events and bounce back ^15, 16^. Resilience is broadly defined as the remarkable ability to rebound from adverse emotional experiences by flexibly adapting to challenging circumstances ^17, 18^. Scholars have suggested that resilience comprises protective factors that modify, alleviate, or transform an individual’s response to environmental threats that may lead to negative outcomes ^19–21^. Recent progress in resilience studies has shifted the focus from simply describing individual traits to exploring the intricate interactions between individuals and their dynamic personal, community, and cultural contexts ^5, 22–24^. One model that offers insight into resilient responses is Li’s Multilevel Resilience Framework ^25^. This model proposes that the presence or absence of individual, community, societal, and systemic resources influences resilient outcomes. Initially designed to explain the resilience of individuals living with HIV, it has evolved into a comprehensive framework for researchers and a practical tool for advocates and communities to comprehend multilevel resilience sources and take targeted action to address them. The development and demonstration of resilience may come more naturally to some individuals than others. It is widely acknowledged that resilience can be cultivated ^22^, and therefore, it is imperative to consider universal interventions aimed at nurturing resilience to uphold mental well-being throughout the challenging times brought on by the pandemic.

Projections suggest that we may need to coexist with COVID-19 for several years, emphasizing the importance of addressing the psychological resilience in mitigating and preventing the negative impacts of the COVID-19 pandemic ^26^. Given the unique challenges posed by COVID-19, such as prolonged and intermittent lockdowns, social isolation, and a global economic downturn, it is crucial to gain a better understanding of the associated factors that contribute to psychological resilience. This understanding will allow us to provide better guidance to policymakers, regulators, and public health officials on supporting and promoting resilience and well-being in future public health crises.

Despite the heightened interest in the concept of resilience and its practical applications, no systematic review has been conducted to comprehensively analyze and synthesize the findings from existing literature on psychological resilience during COVID. Previous systematic reviews related to resilience and the COVID-19 pandemic have looked at the effects of the pandemic on a specific population, including healthcare providers ^27–30^, young generation ^31, 32^, or specific countries ^33–35^, lacking a comprehensive approach across all demographics. Some studies focused on resilience at individual mental health strategies ^26^, psychological intervention ^36^, or neurobiological and developmental aspects ^5^ but overlook broader community and structural levels and biopsychosocial associated factors. There is a great need for a comprehensive review of global literature, especially those published from 2020 to 2024, capturing more recent data and insights from the later stages of the pandemic. This global and cross-disciplinary approach will allow for a broader and more current understanding of resilience across different age groups, occupations, and countries and can provide a more nuanced understanding of resilience during COVID-19, addressing gaps in the existing reviews by exploring how different contextual factors influence resilience outcomes. Therefore, in our systematic review of current literature on psychological resilience during the COVID-19 pandemic, our objectives were to identify key factors contributing to psychological resilience during this period and to extract insights that can be applied to future preparedness. Our research questions are as follows:

- How has psychological resilience been defined and measured?
- What are the factors associated with psychological resilience?
- What interventions have been used to increase psychological resilience during the COVID-19 pandemic, and how can these strategies be adapted for future crises?

## 2. METHODS

### 2.1. Study design

This systematic review adhered to the PRISMA preferred items for systematic reviews ^37^, ensuring the study’s credibility and reliability. The protocol was pre-registered on the Prospero website with ID number CRD42024590489.

### 2.2. Data Sources

Four major electronic bibliographic databases were searched in February 2024 by two authors (AP, JN), including PubMed, PsycINFO, ScienceDirect, and Web of Science. These databases were selected to ensure comprehensive coverage across biomedical, psychological, and social science disciplines. Preliminary searches were conducted to refine the queries and ensure relevance. The search was conducted using a combination of keywords and Medical Subject Headings (MeSH) terms related to “psychological resilience” and “COVID-19.” (**Supplementary Table 1**) The search strategy was tailored to each database, and search results were filtered to include only peer-reviewed articles published in English between 2020 and 2024. Additionally, we manually checked the reference lists of included articles and relevant review papers to identify additional studies that were not revealed during the formal database search.

### 2.3. Inclusion and Exclusion Criteria

The eligibility criteria for this review were defined to include studies that specifically focused on psychological resilience during the COVID-19 pandemic. The inclusion criteria were limited to quantitative research articles published in peer-reviewed journals between 2020 and 2024, written in English, and conducted on human participants. Studies that investigated psychological resilience, its associated factors, and interventions aimed at enhancing resilience during the pandemic were considered relevant.

Exclusion criteria were applied to studies that focused primarily on resilience at other levels (e.g., organizational, structural). Qualitative studies, commentaries, editorials, review articles, or theoretical papers were excluded. Papers that were not available in English or were based on non-human research were also excluded from the review.

### 2.4. Screening Process

All initial search results were imported into RAYYAN, a reference management software, which was used to organize and facilitate the screening process. Two researchers (AP, NJ) independently conducted the initial screening based on titles and abstracts. Studies that did not meet the inclusion criteria were excluded at this stage. Following the title and abstract screening, the full texts of potentially relevant articles were retrieved and reviewed for eligibility based on the pre-established inclusion and exclusion criteria. Any discrepancies were resolved through discussion. In cases where the agreement could not be reached, a third researcher (SQ) was involved to adjudicate. This multi-stage review process ensured that all eligible studies were systematically identified and included in the final analysis.

### 2.5. Data Extraction

Data extraction was carried out using standardized data extraction forms designed to capture key study characteristics and findings. For each included study, the following information was extracted: author(s), year of publication, study design, sample size, population characteristics (e.g., age, gender, marital status), the resilience definition and measurement tools used, key findings and associated factors related to psychological resilience, any reported interventions aimed at fostering resilience during the COVID-19 pandemic, limitation and future suggestion of included studies. Two independent researchers (NJ, AP) extracted data from the included studies. All extracted data were recorded in a shared database to ensure consistency and accuracy. Any discrepancies in data extraction were resolved through discussion among the researchers. The extracted data was used to inform the synthesis of the review and to identify common themes across studies.

### 2.6. Quality Assessment

Two independent coders (AI and JN) assessed the methodological quality of our included articles. The third assessor (AP) addressed disagreements between the two. This study used three quality assessment tools produced by the US National Institutes of Health (NIH)^38^ for the different study designs of the included studies; the Quality Assessment Tool for Observational Cohort and Cross-Sectional Studies, the Quality Assessment Tool for Controlled Intervention Studies, and Quality Assessment Tool for Before-After (Pre-Post) Studies With No Control Group.

All included papers were assessed along each question/dimension of the NIH tools as “Yes,” “No,” “Not Reported (NR),” “Cannot Determine (CD),” or “Not Applicable (NA).” The “NR” option was used for studies that could potentially be answered “Yes” for the question but did not report the required information; for instance, question 3 in the cross-sectional tool asks about the participation rate of eligible persons if a study only reports the number of participants but not the number of all of those who were approached initially this question would be “NR.” The “CD” option was for those questions that the text’s information was vague or inadequate to answer because of lack of clarity. The “NA” option was used when one of the instrument’s criteria could not be evaluated due to the type of study (such as a cross-sectional design).

According to the NIH tool guidelines, for cross-sectional studies, D6 (Exposure assessed before outcome measurement) and D7 (Sufficient timeframe to see an effect) were instructed to receive “No”. Additionally, D10 (Repeated exposure assessment), D12 (Blinding of outcome assessors), and D13 (Follow-up rate) did not apply to cross-sectional studies, so these items were also labeled “NA.” Therefore, the overall quality assessment of these studies was determined based on the sum of “Yes” responses to applicable questions (which were nine questions for cross-sectional studies), intentionally excluding the non-applicable questions from the comprehensive evaluation. Studies with at least six “Yes” responses were rated as “Good,” while those with 4 to 5 were considered “Fair,” and those with 3 or below were considered as Poor.” However, D6, D7, D10, D12, and D13 were considered in the comprehensive evaluation of the included longitudinal studies.

### 2.7. Data Synthesis

A narrative synthesis was conducted to identify common themes, including definitions of psychological resilience, associated factors, and strategies employed to enhance resilience during the pandemic. These themes informed the analysis by organizing and interpreting the extracted data, addressing the review’s research questions, and providing insights into the factors associated with psychological resilience. Additionally, the synthesis guided the categorization of measurement instruments and interventions, facilitating a comprehensive understanding of the methodologies and practices reported in the studies.

## 3. RESULTS

The online search retrieved a total of 2054 papers: PubMed (1042), PsycINFO (19), ScienceDirect (880), and Web of Science (113). After removing 614 duplicated records, 1440 papers were included in the title screening. Finally, 68 papers were included in the review, including 49 cross-sectional studies, 16 longitudinal cohort studies, and 3 intervention studies. The process of the literature search and the selection of papers is shown in **Figure 1**.

**Figure 1.**
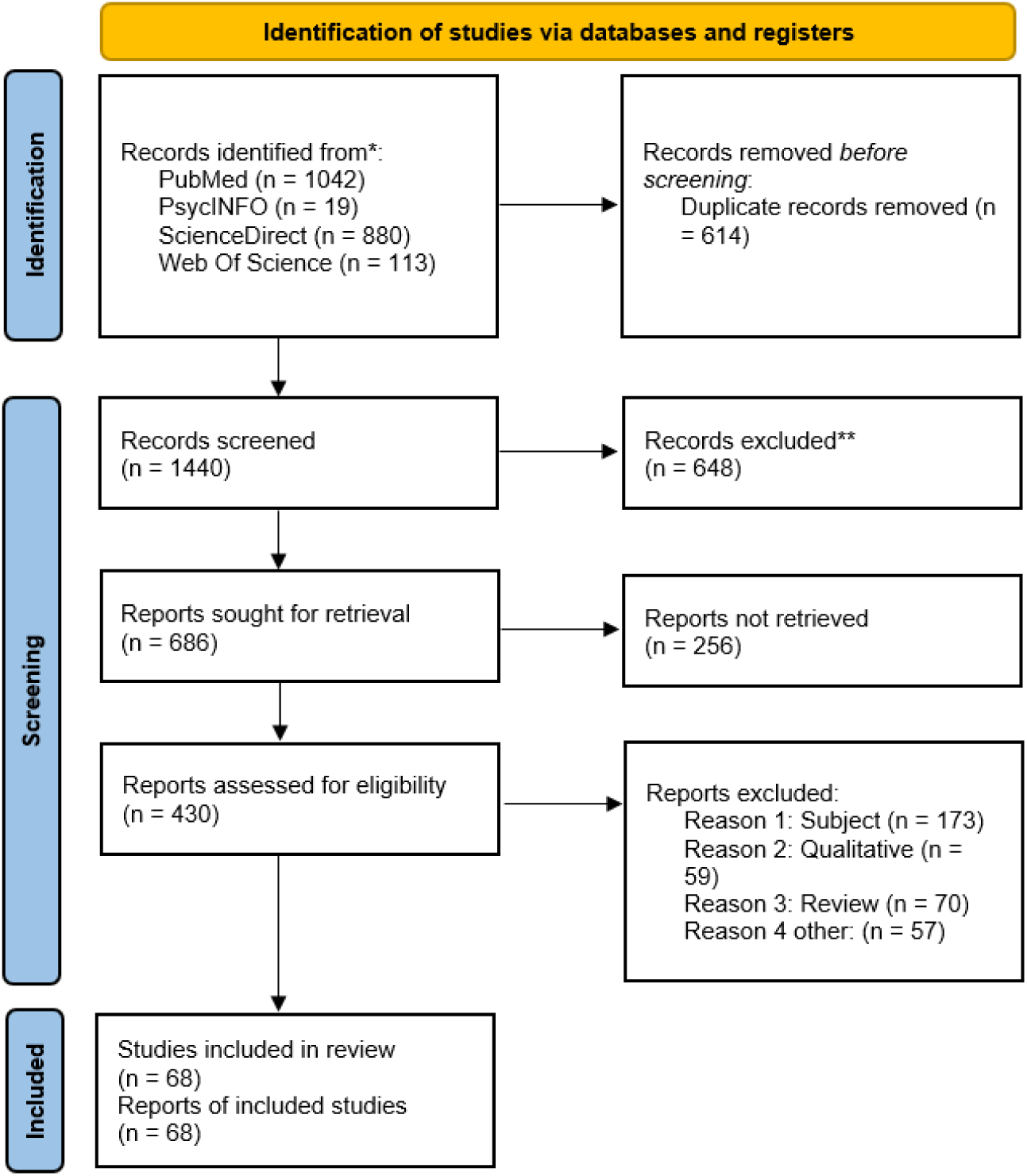
Flow diagram of the literature search and articles selection (adapted from PRISMA 2020 guidelines for systematic reviews).

### 3.1. Study Characteristics

The study characteristics of the included studies are presented in **Table 1**. The geographical distribution of studies in this review highlighted significant regional representation, with Asia, North America, and Europe being the most represented regions. China was the most represented country, contributing 13 studies, followed by the United States and Italy, each with 6 studies. Other countries with multiple representations included Turkey (n=3), Israel (n=3), Brazil (n=2), Australia (n=2), Spain (n=2), Germany (n=2), India (n=2), and Qatar (n=2). A number of countries, including England, South Africa, Greece, and Poland, were represented by a single study each. Six studies focused on multiple countries, with the number of countries represented ranging from 2 to 63.

**Table 1.**
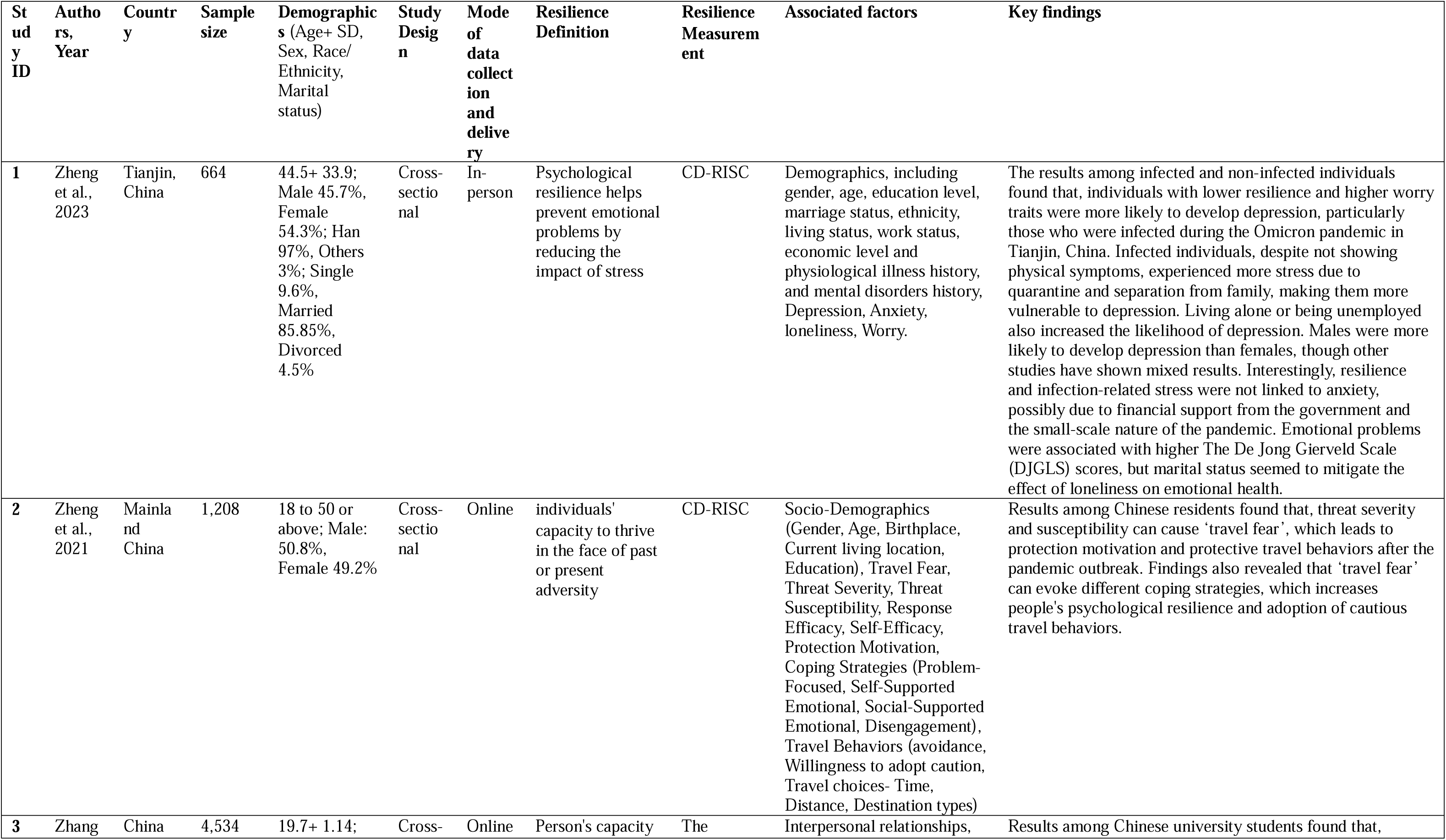
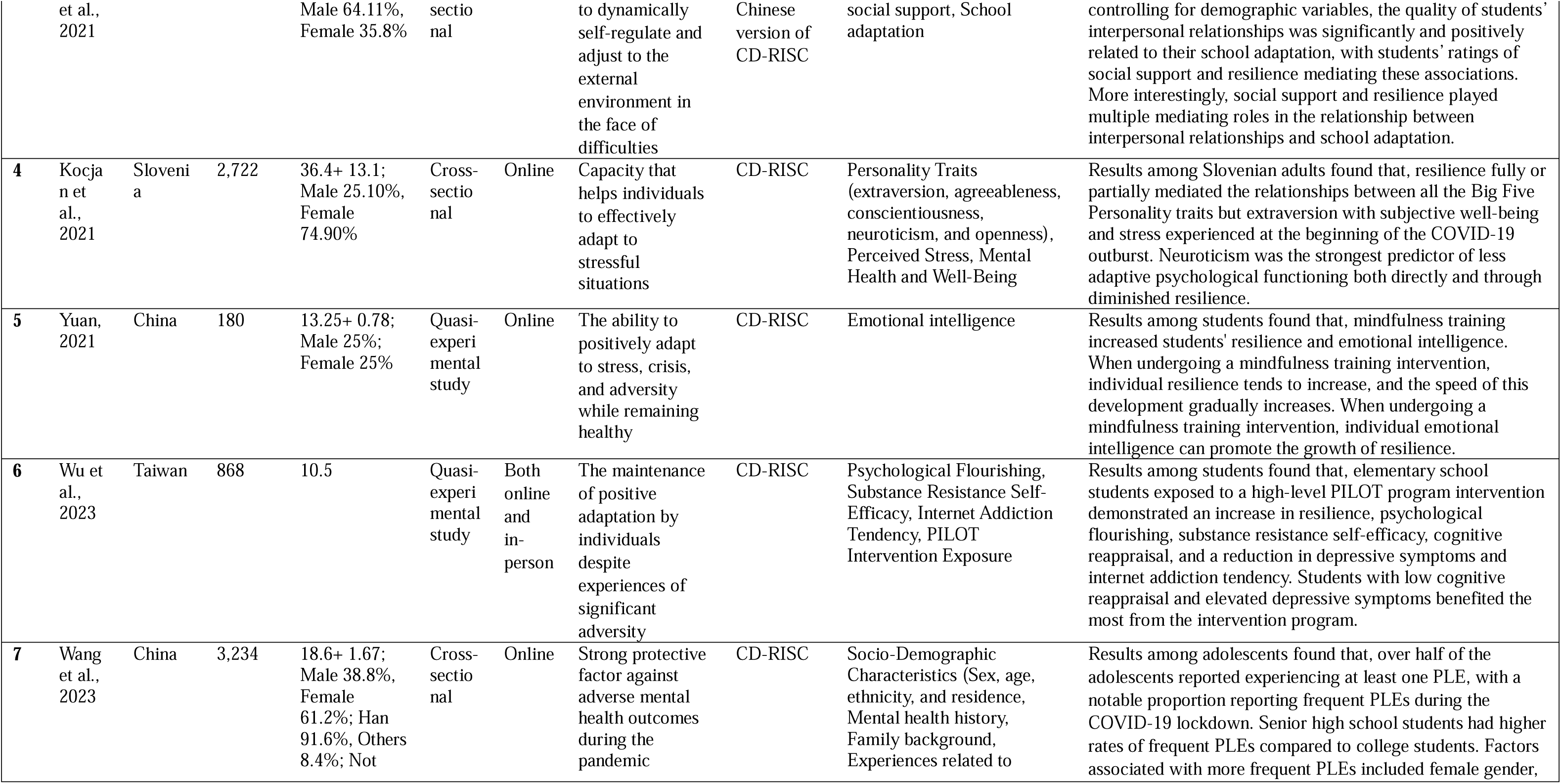
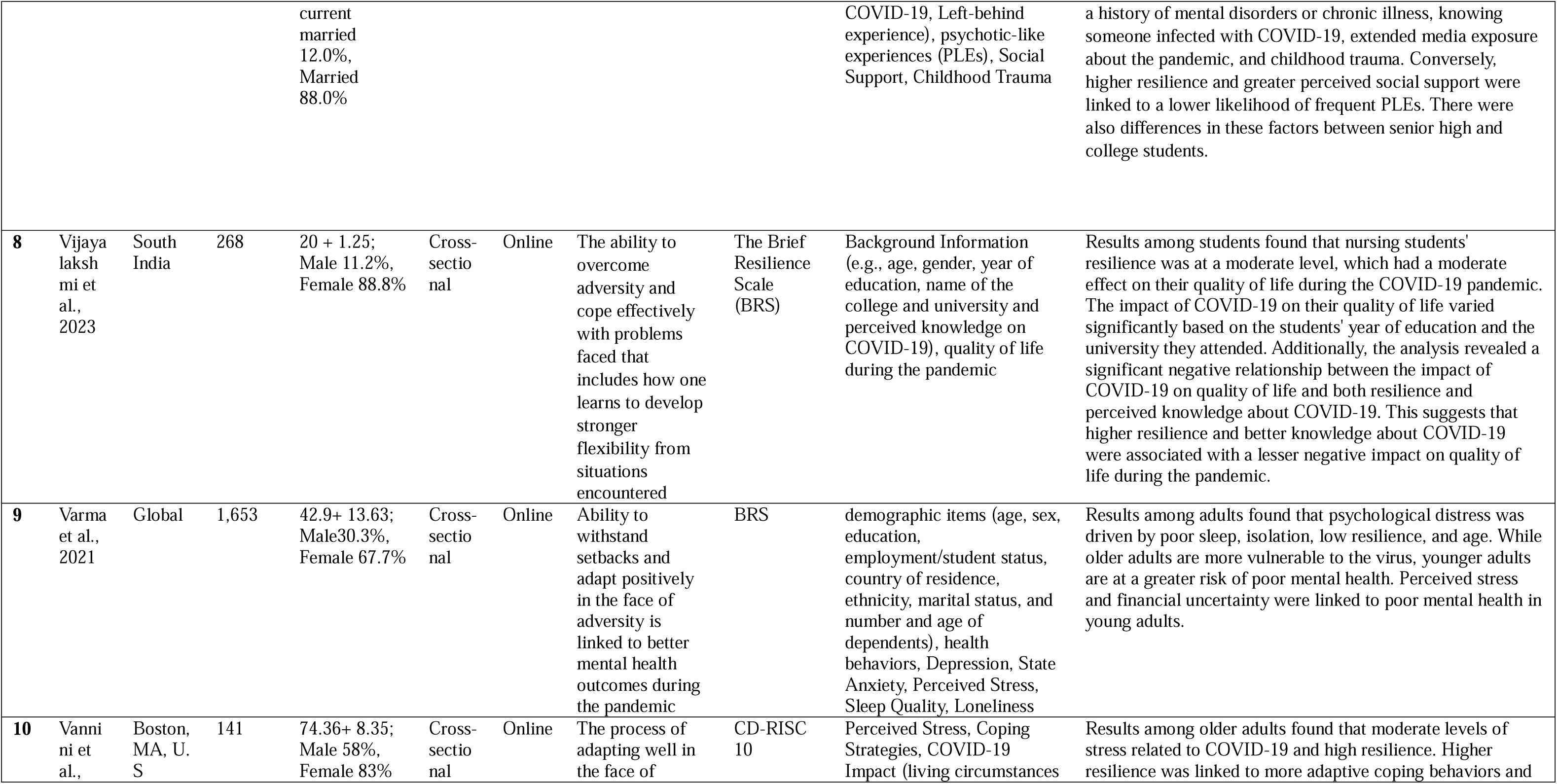
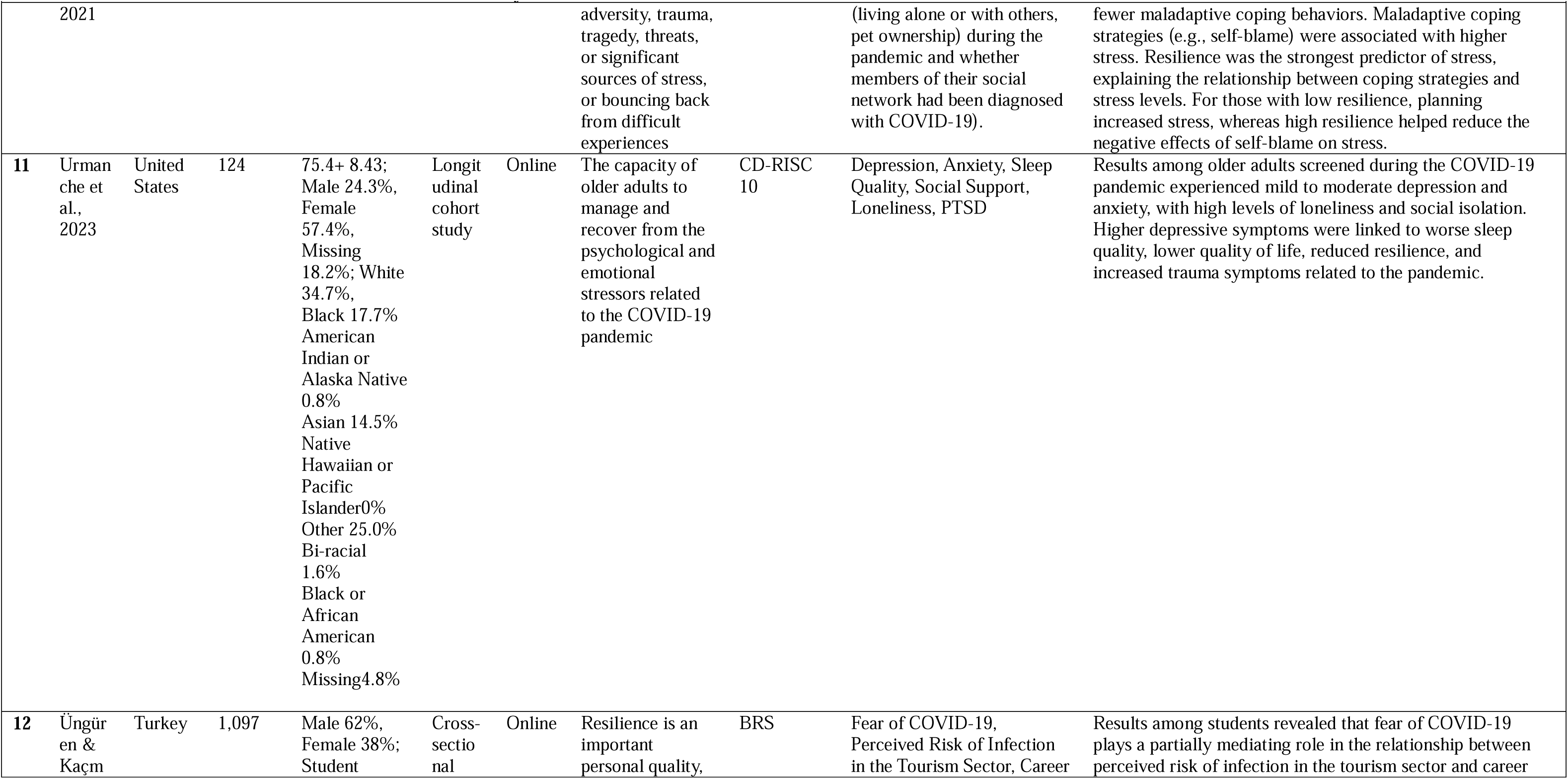
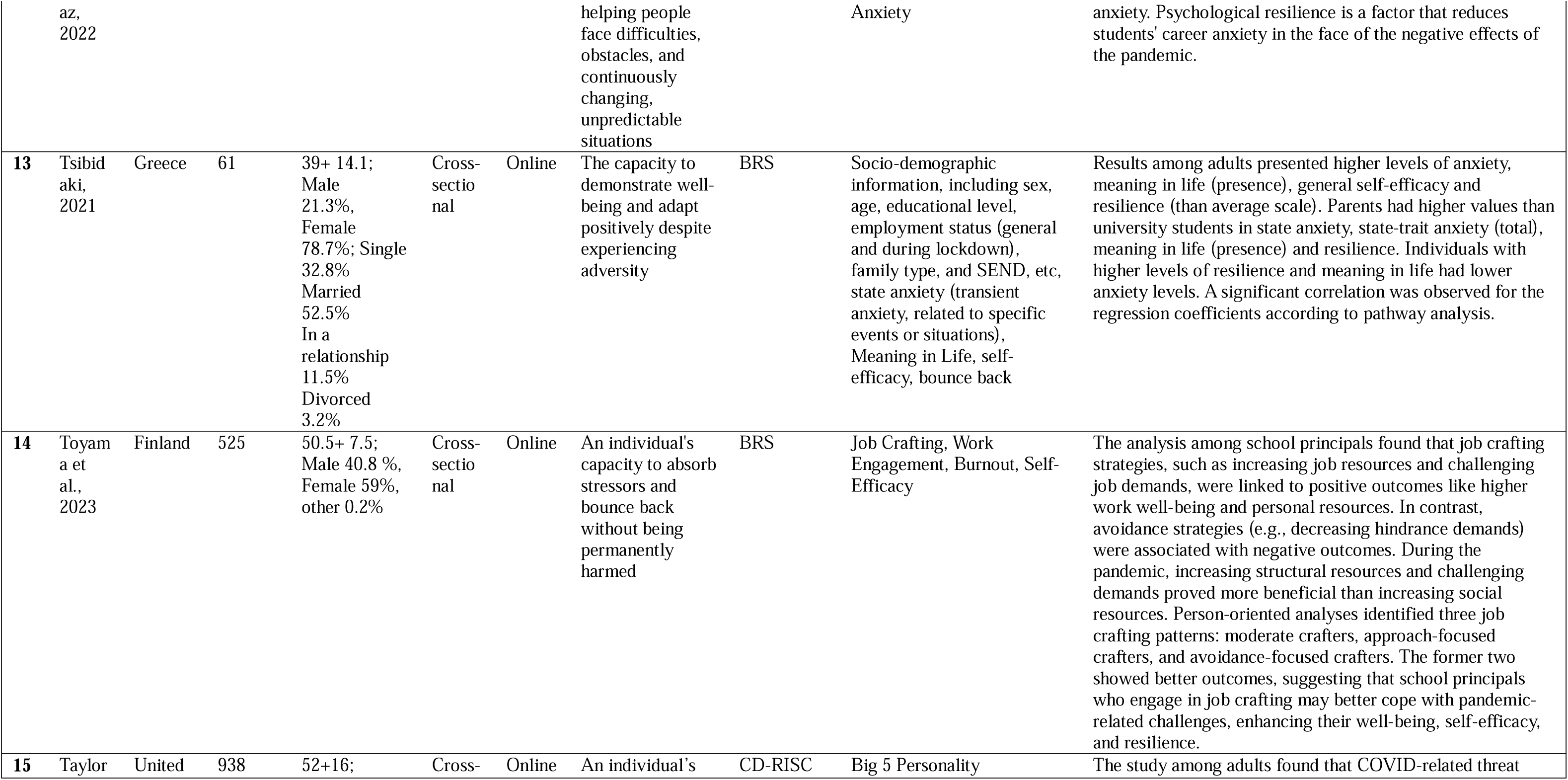
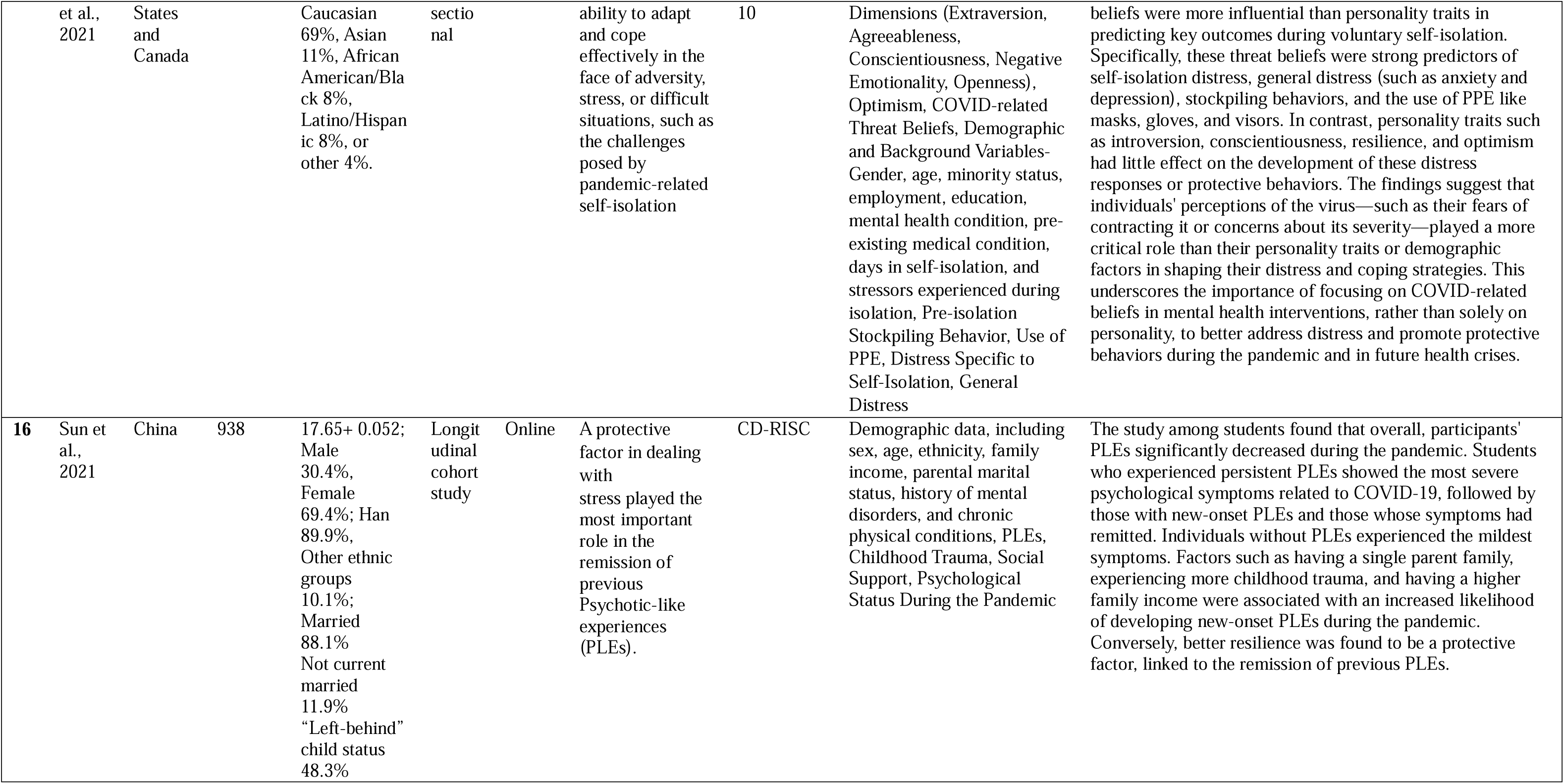
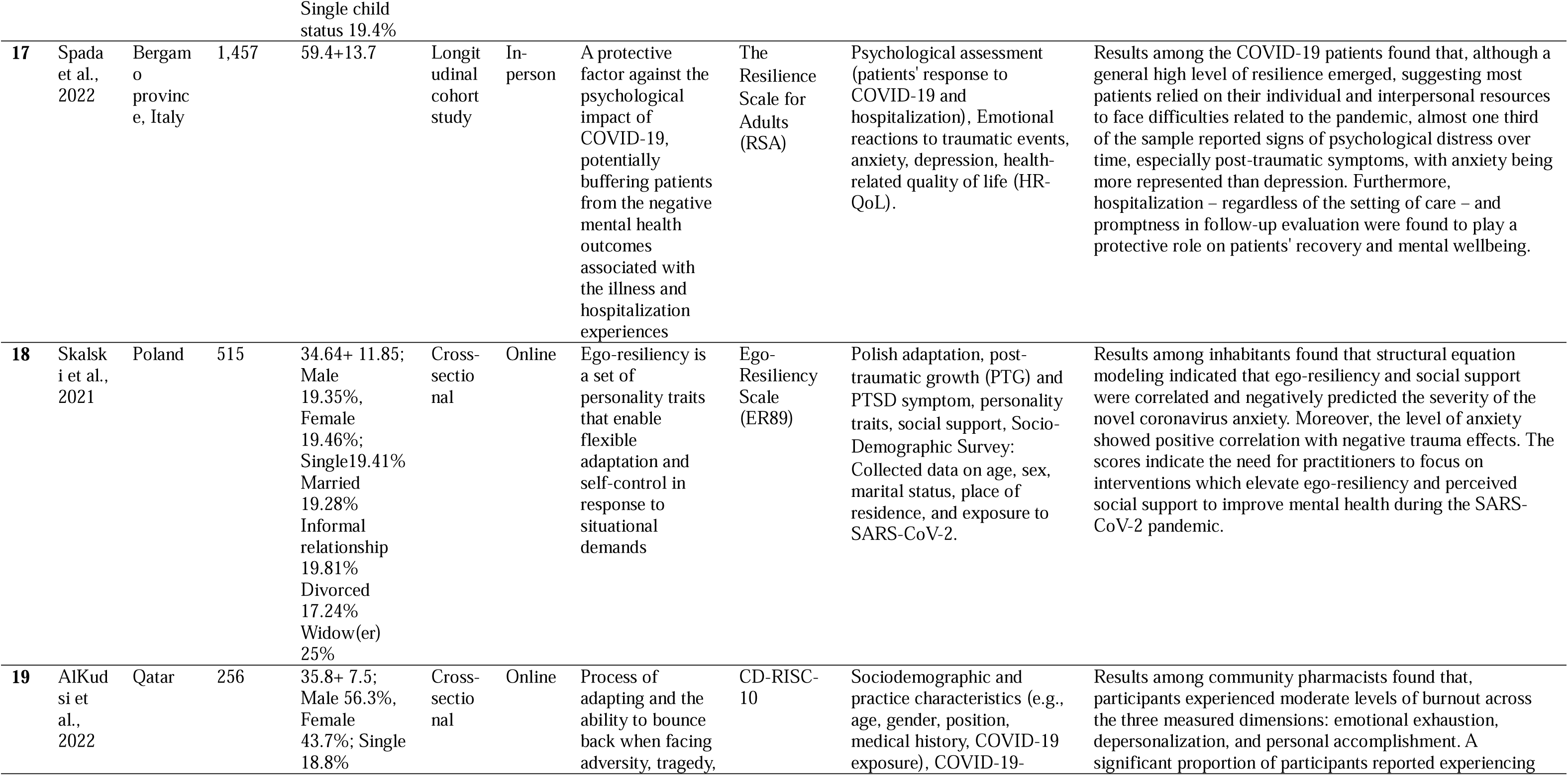
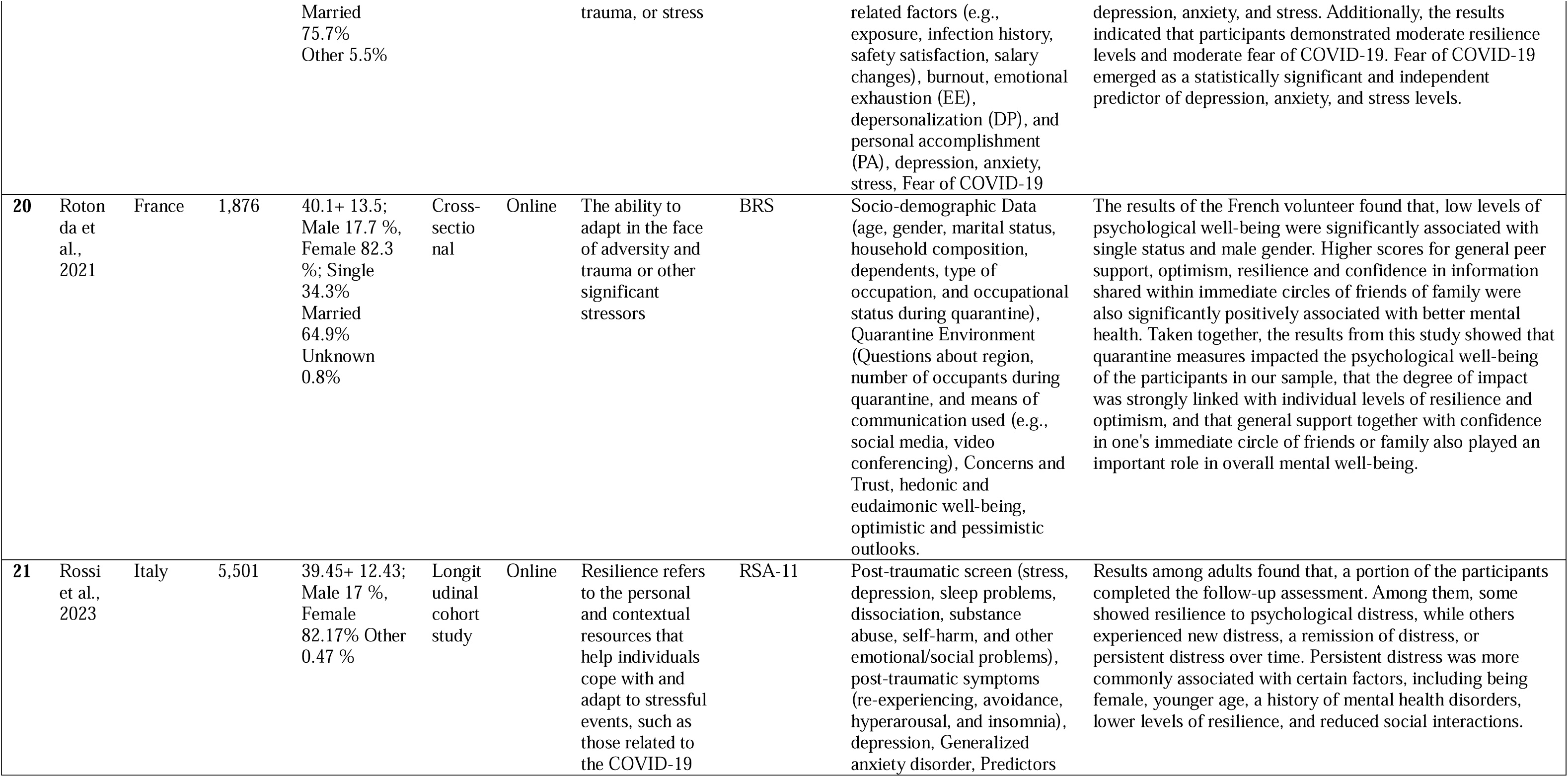
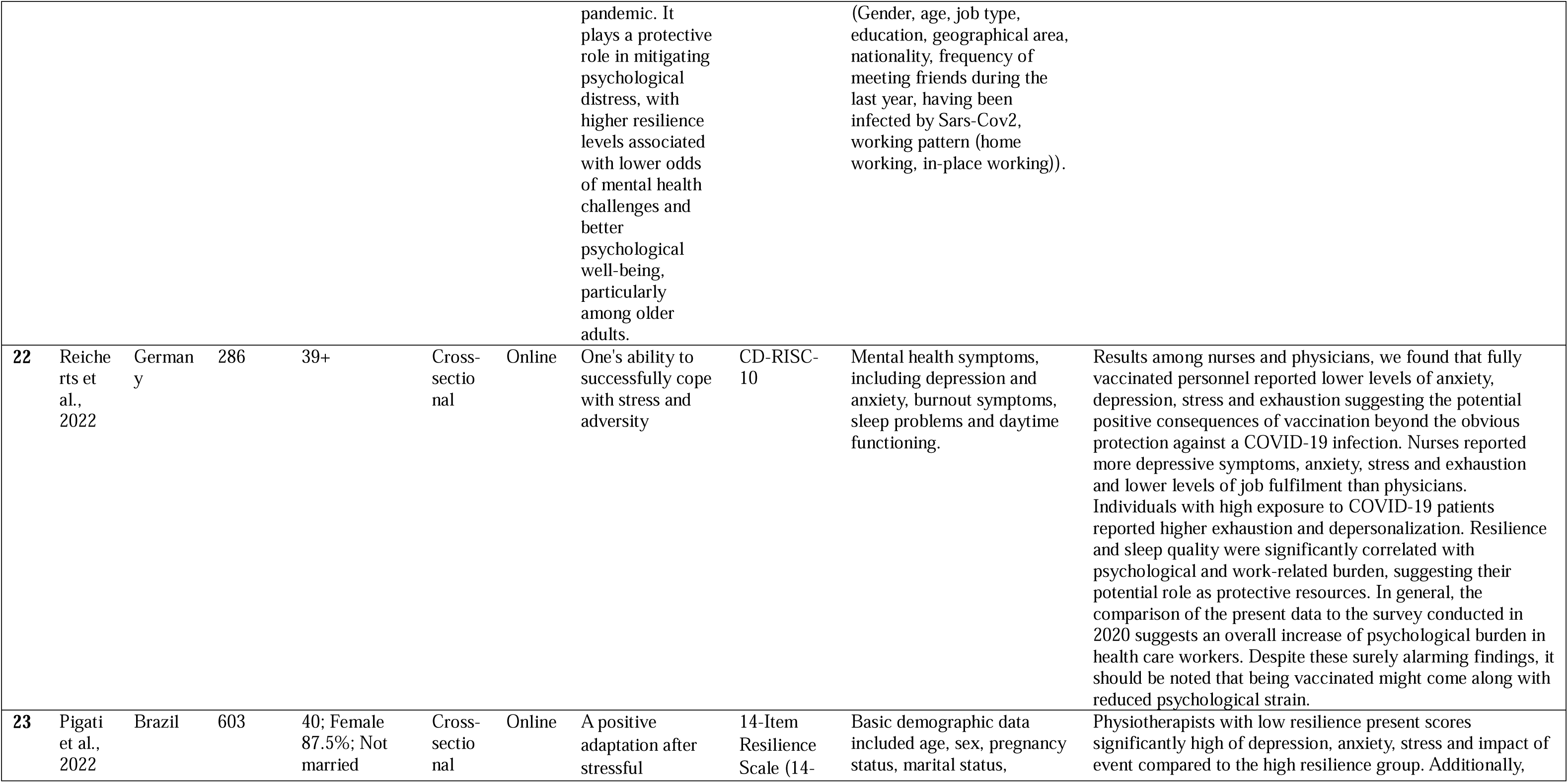
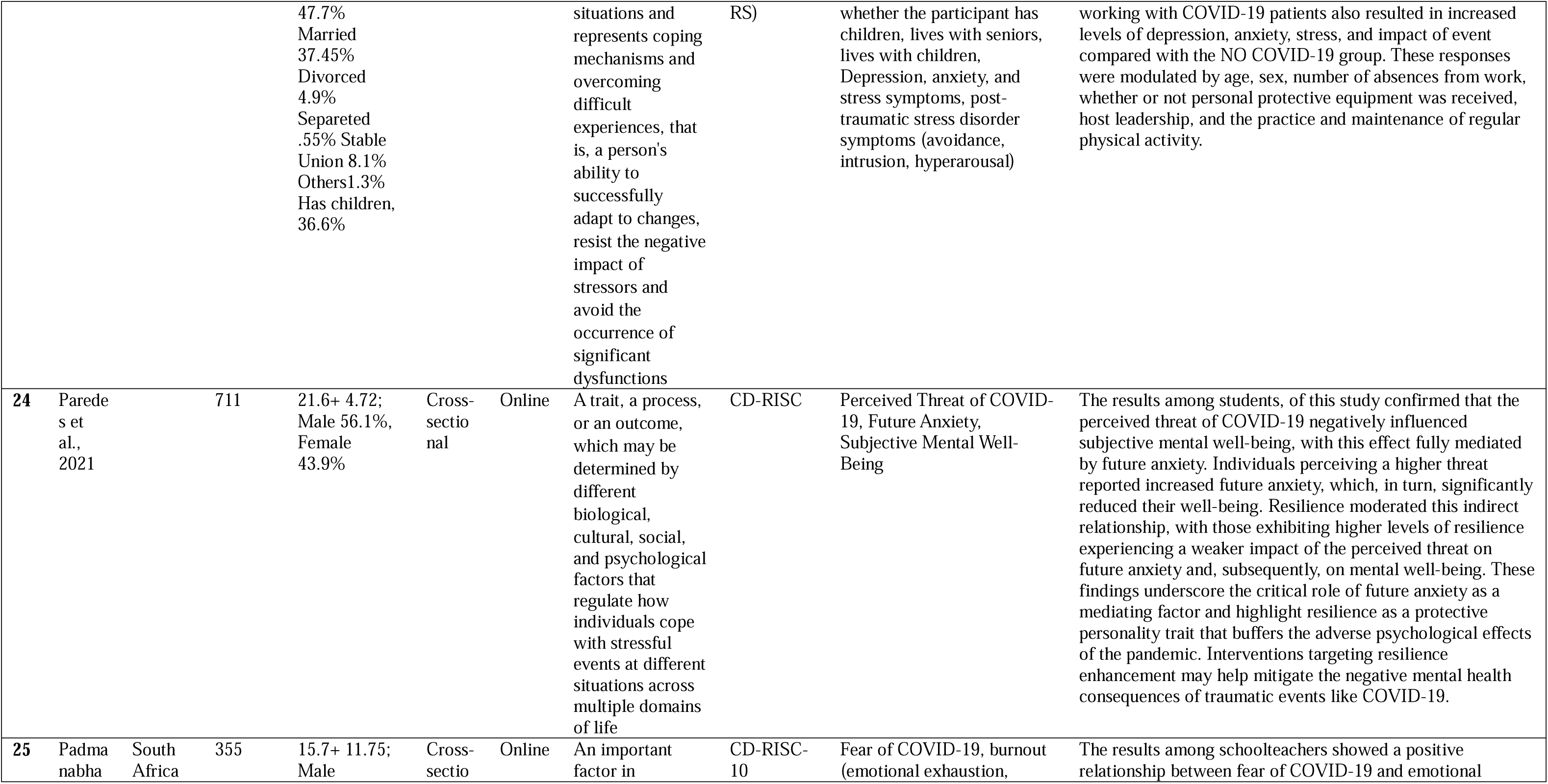
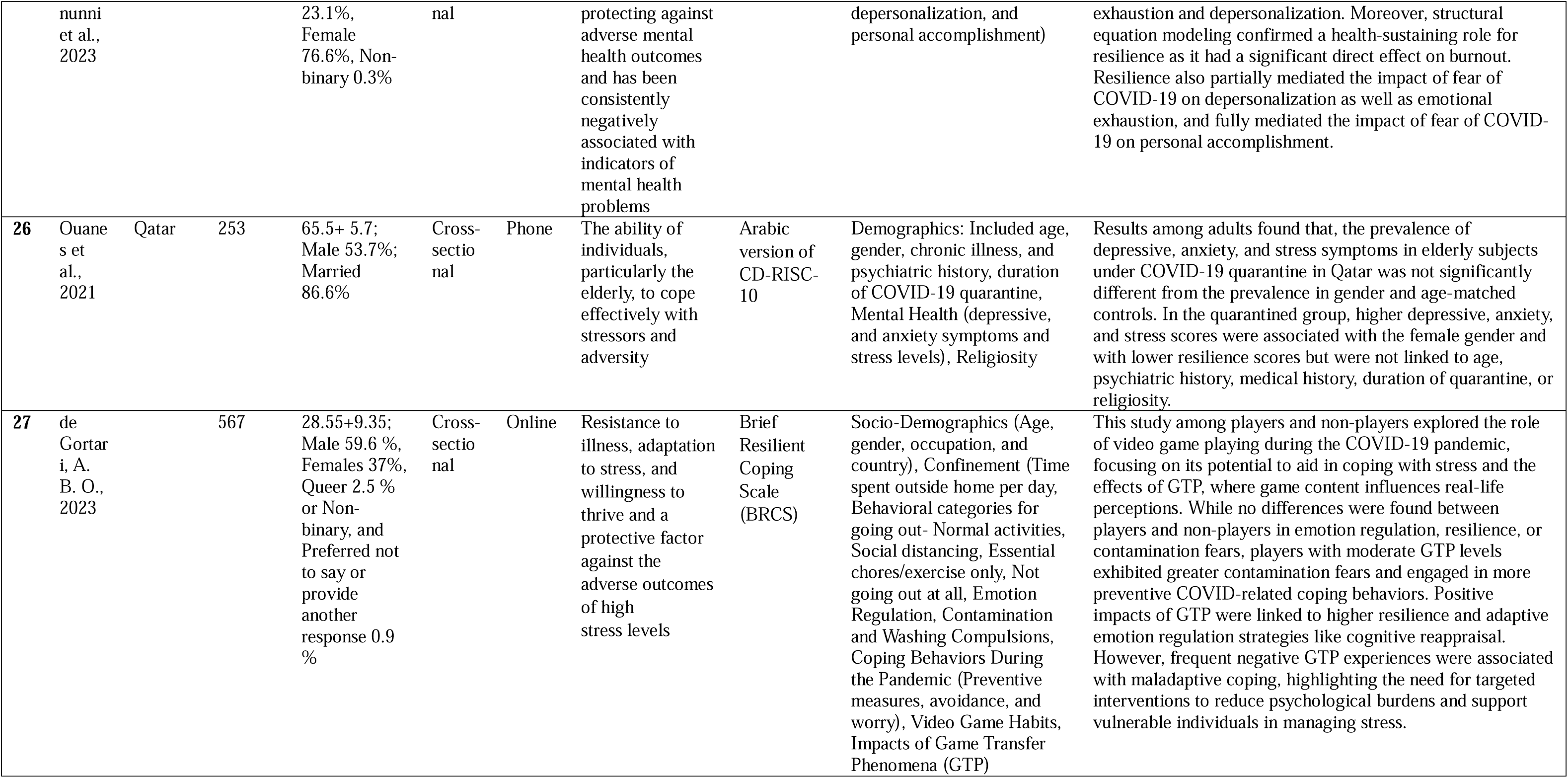
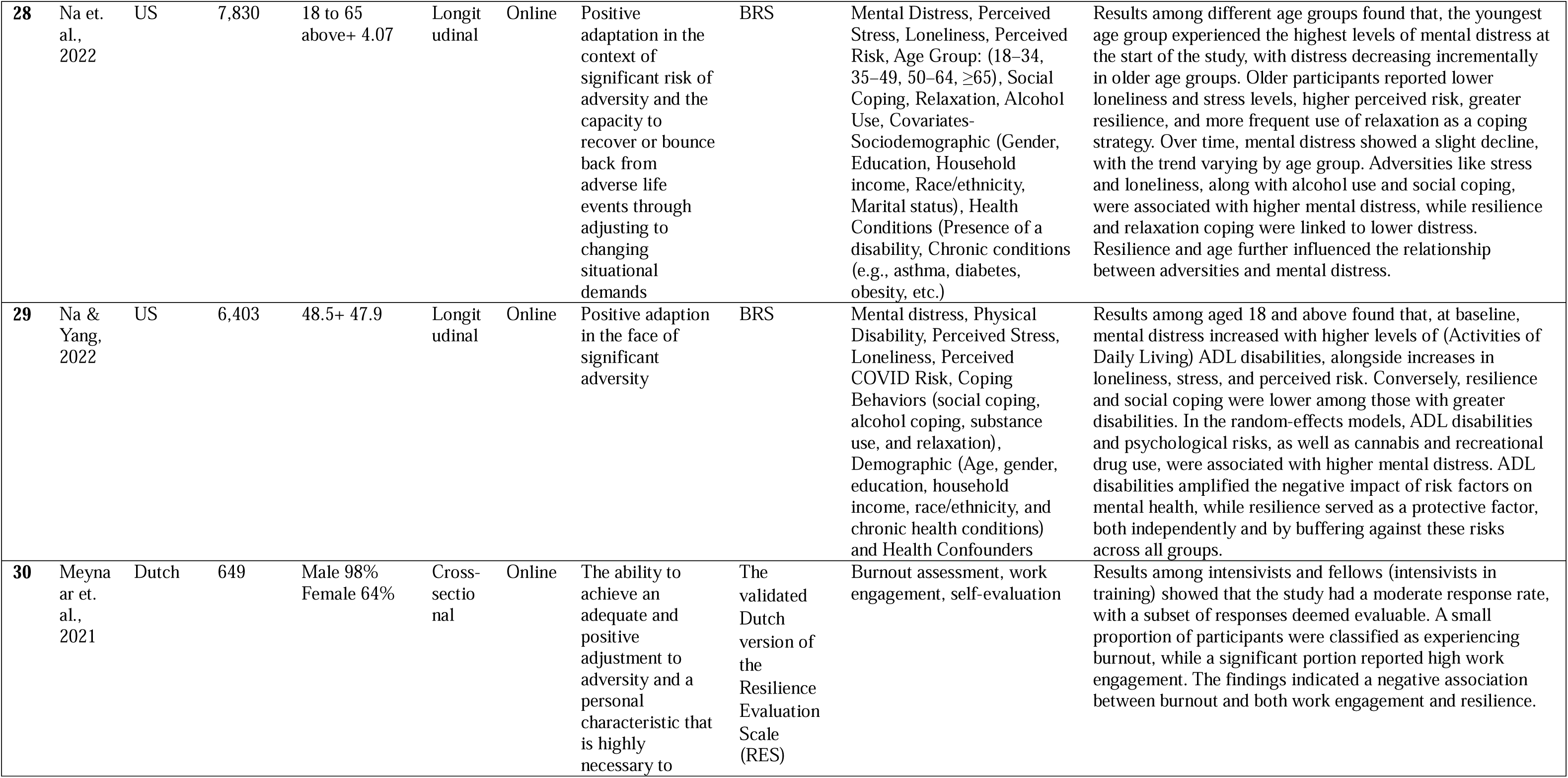
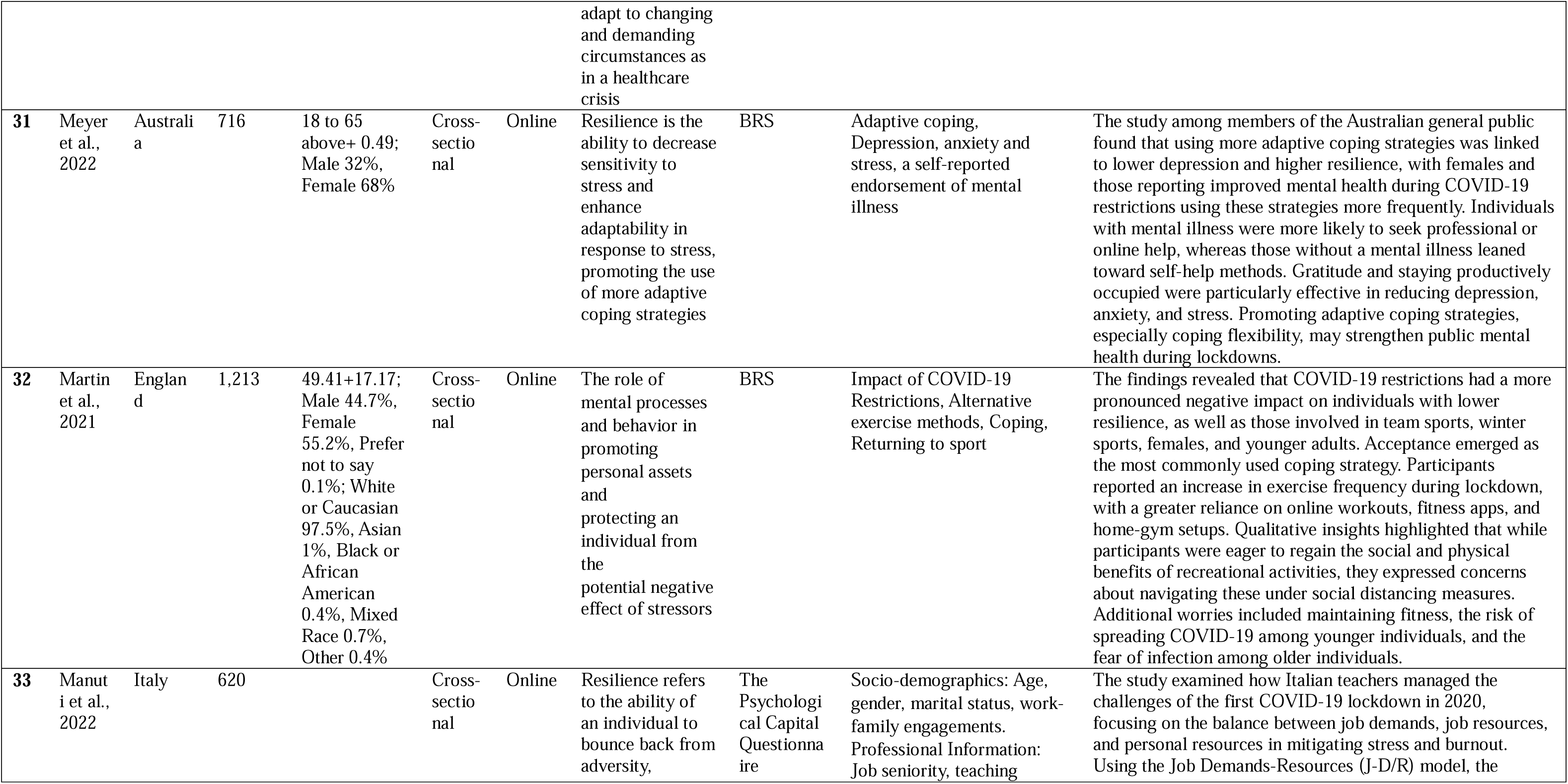
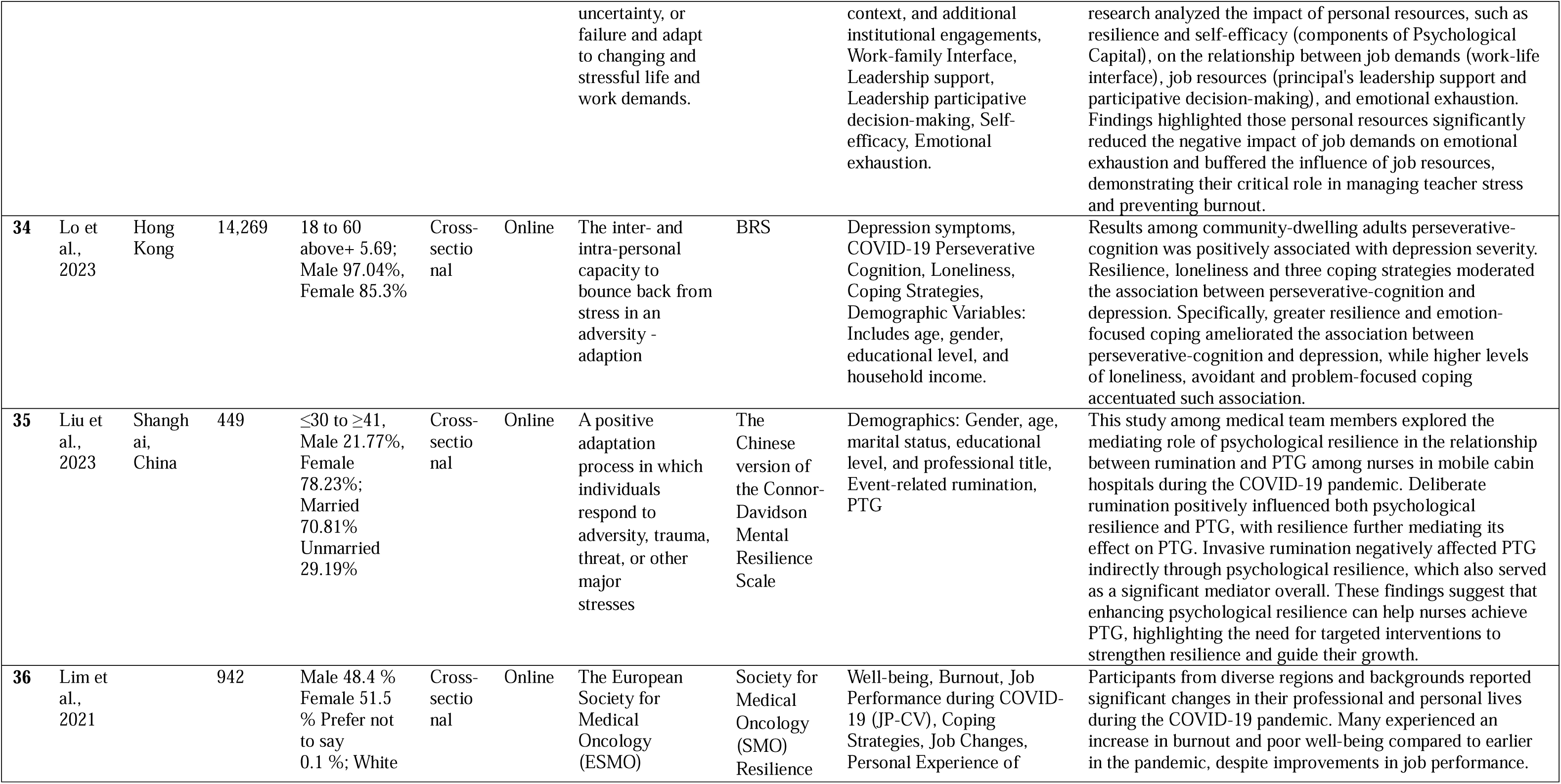
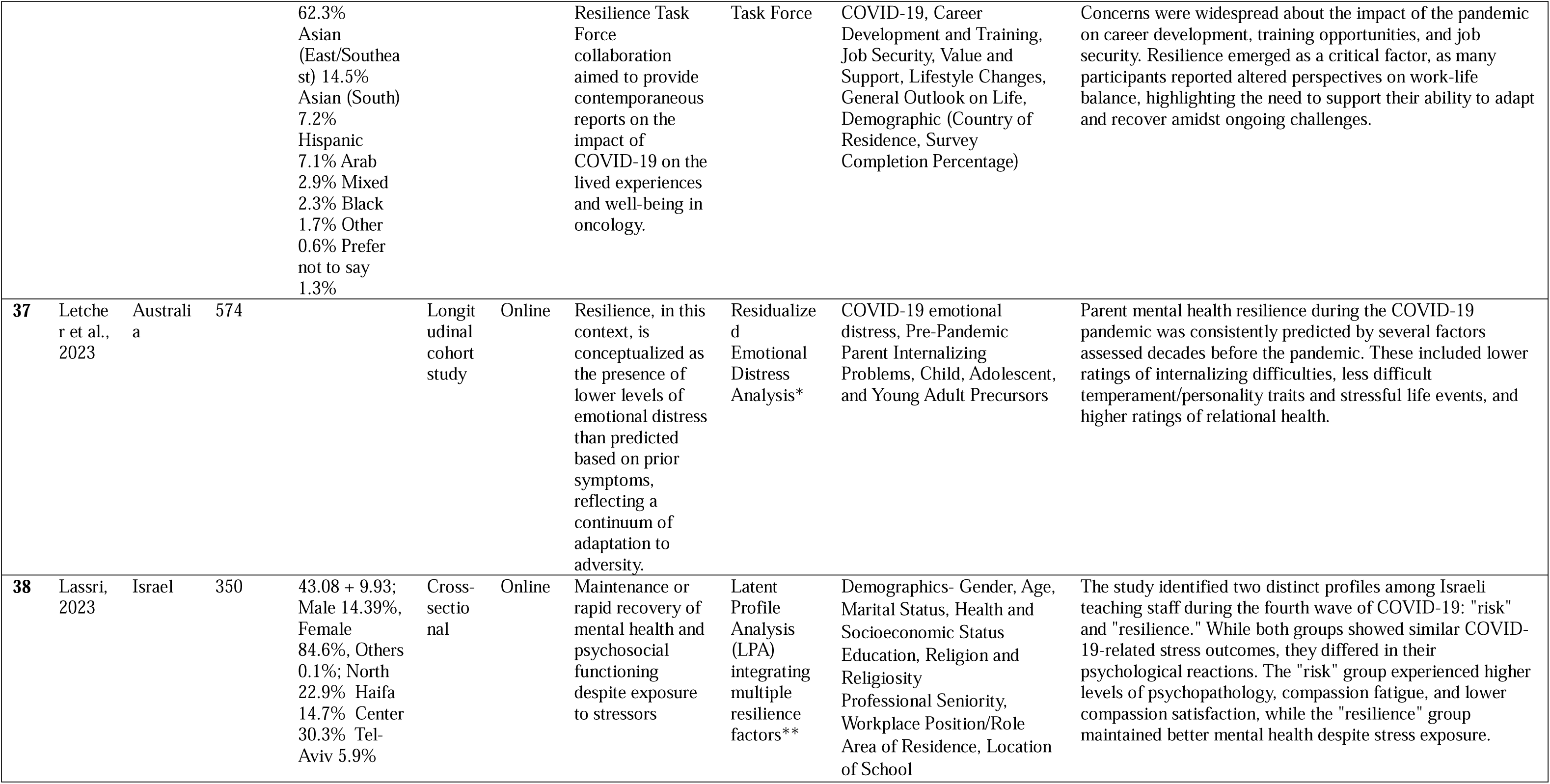
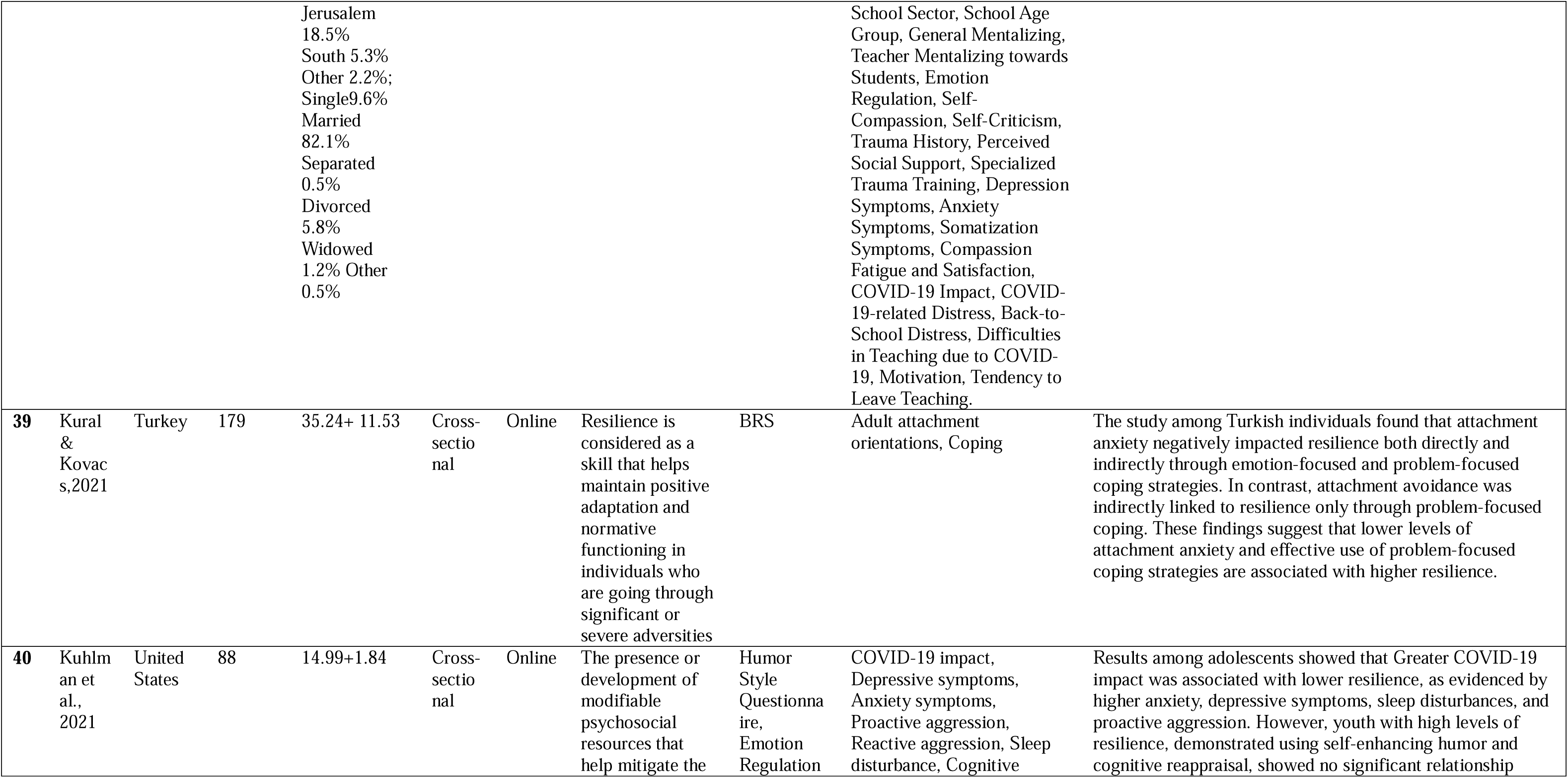
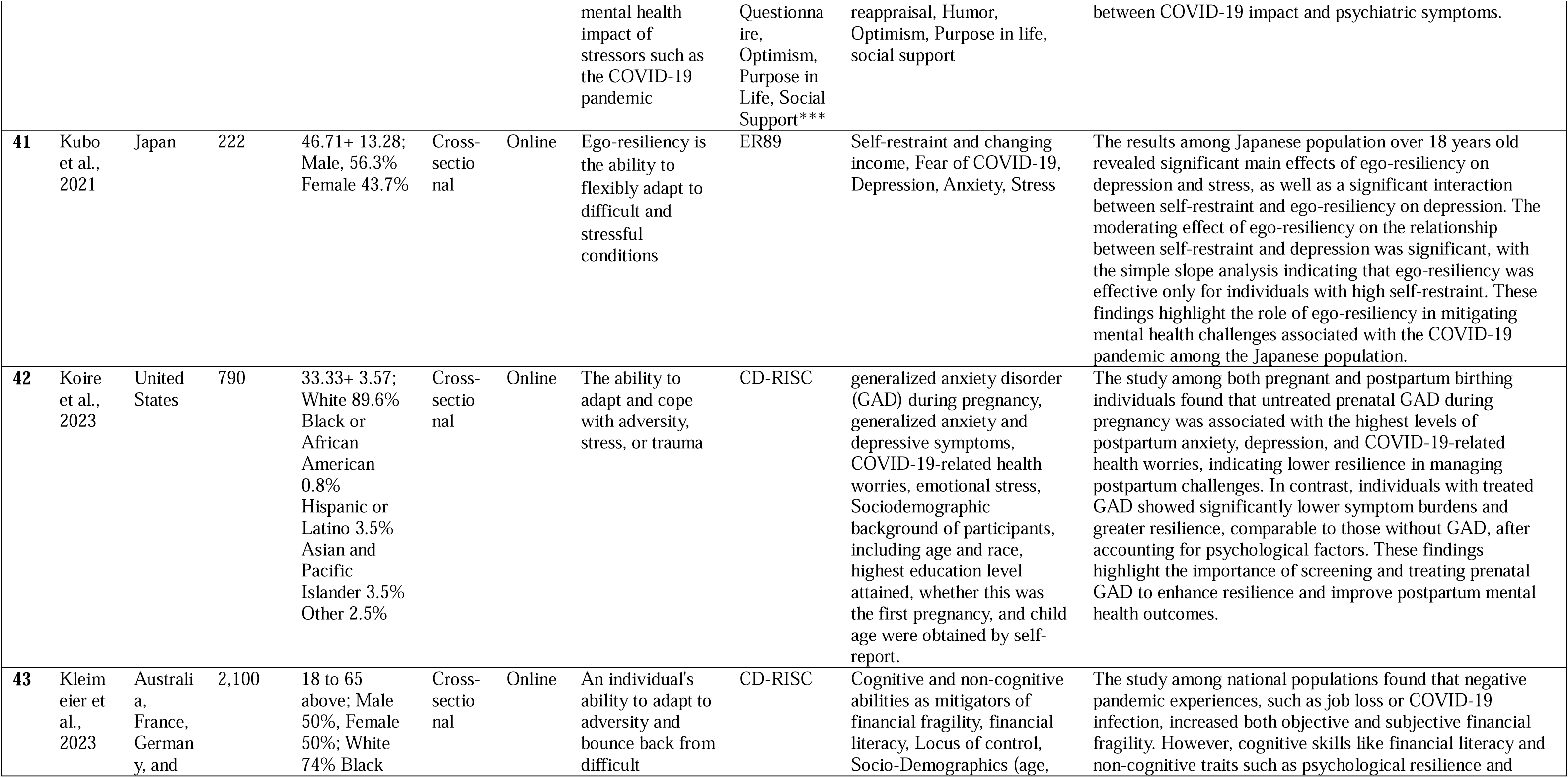
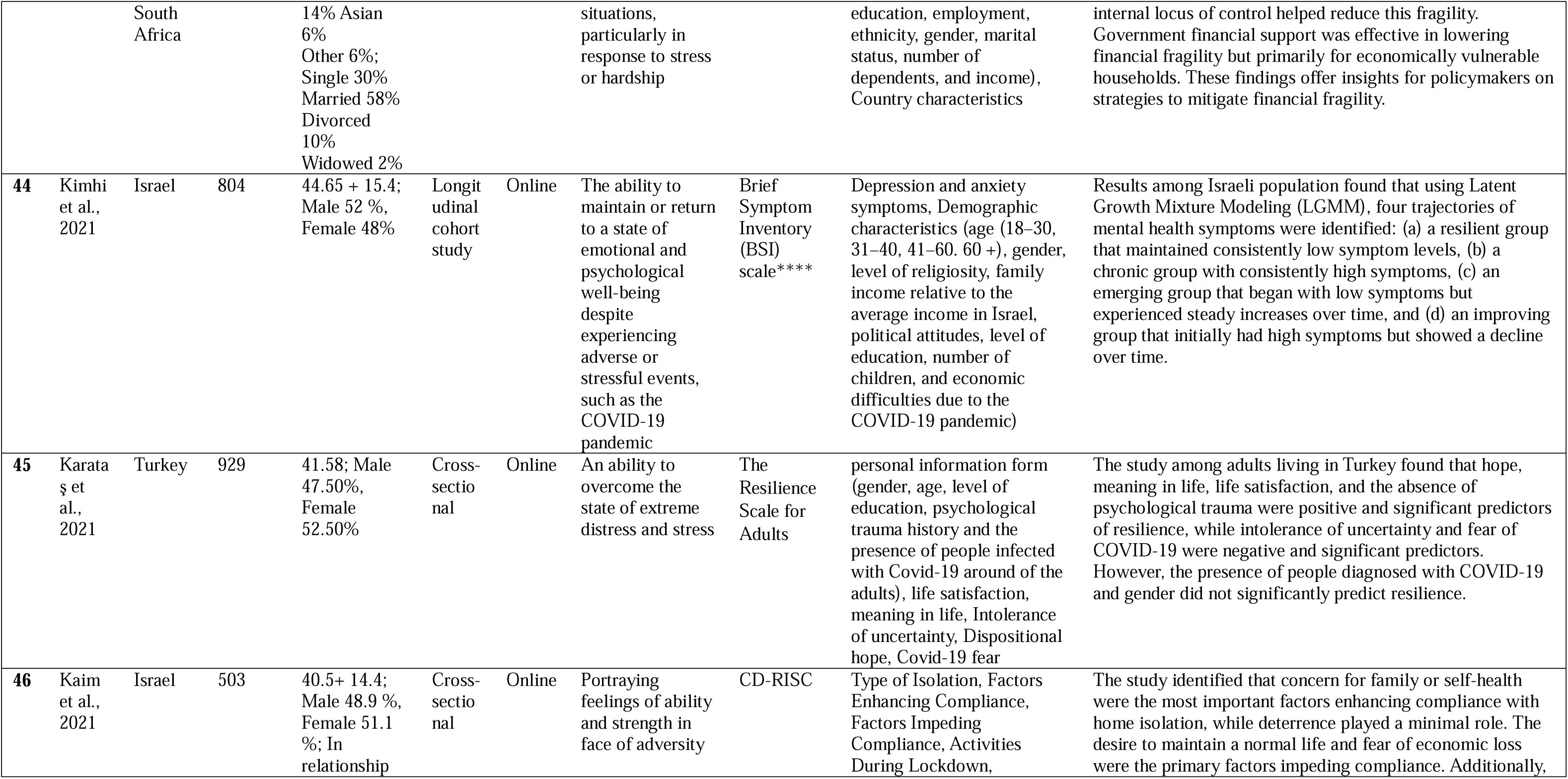
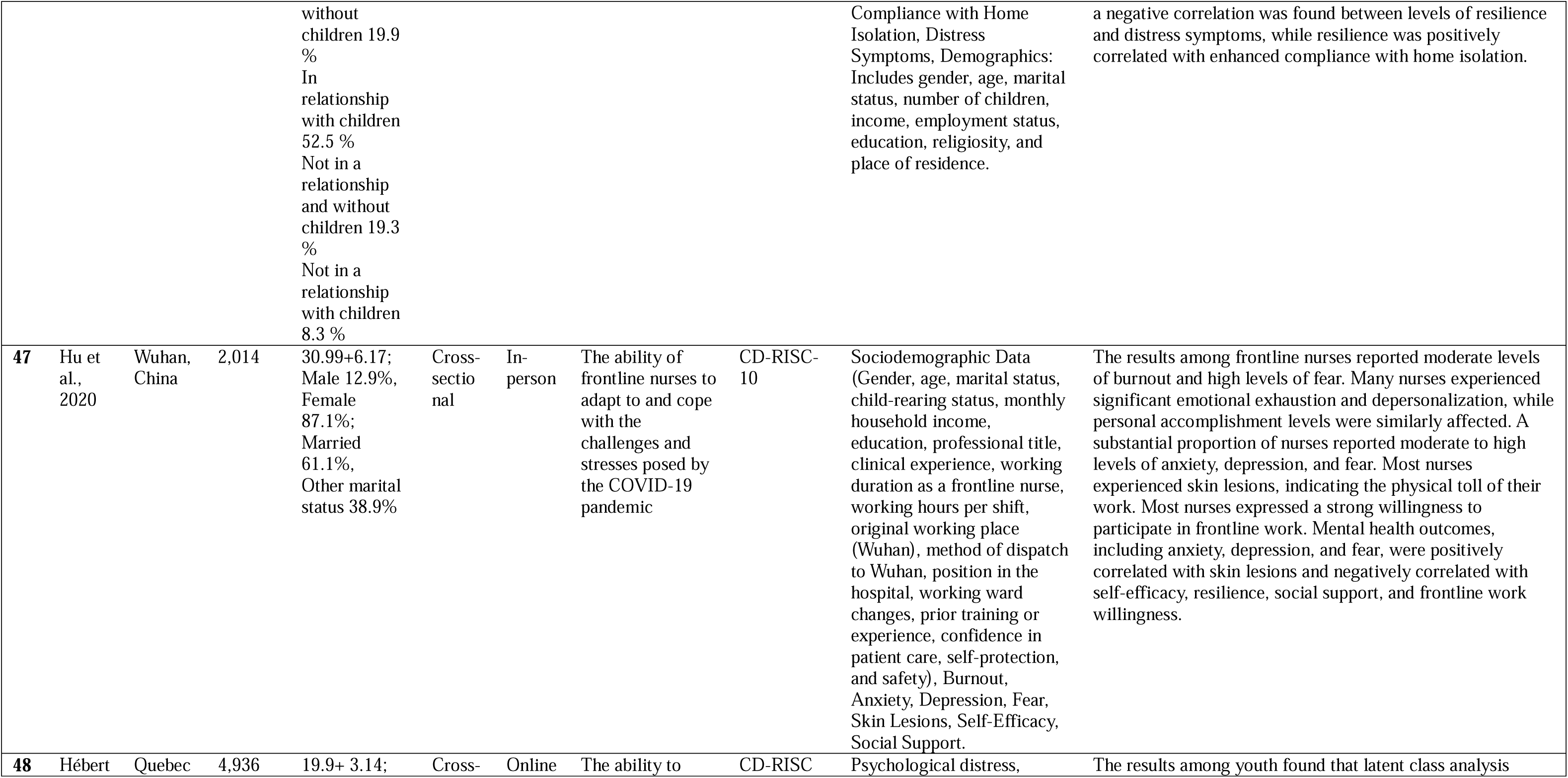
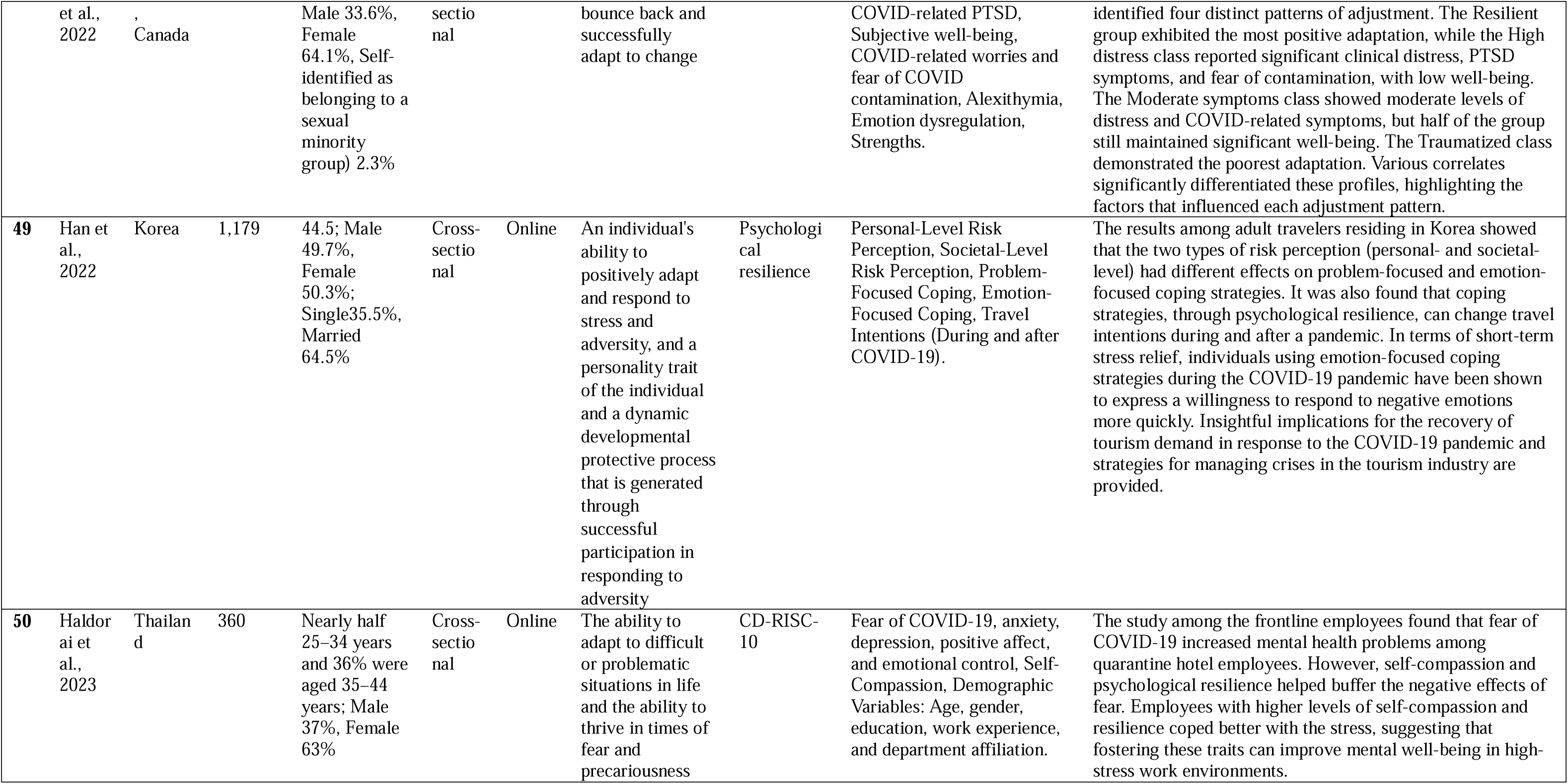
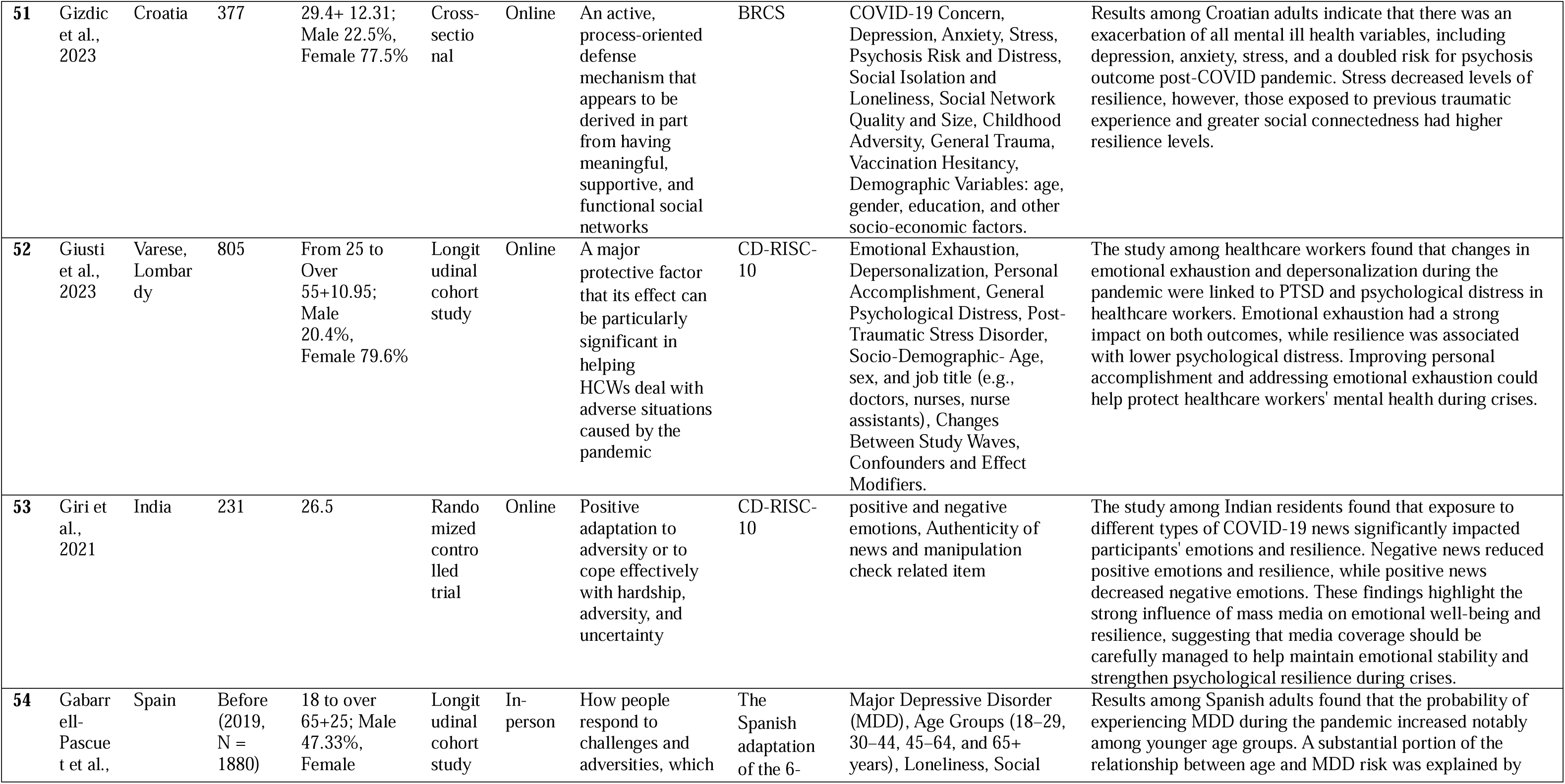
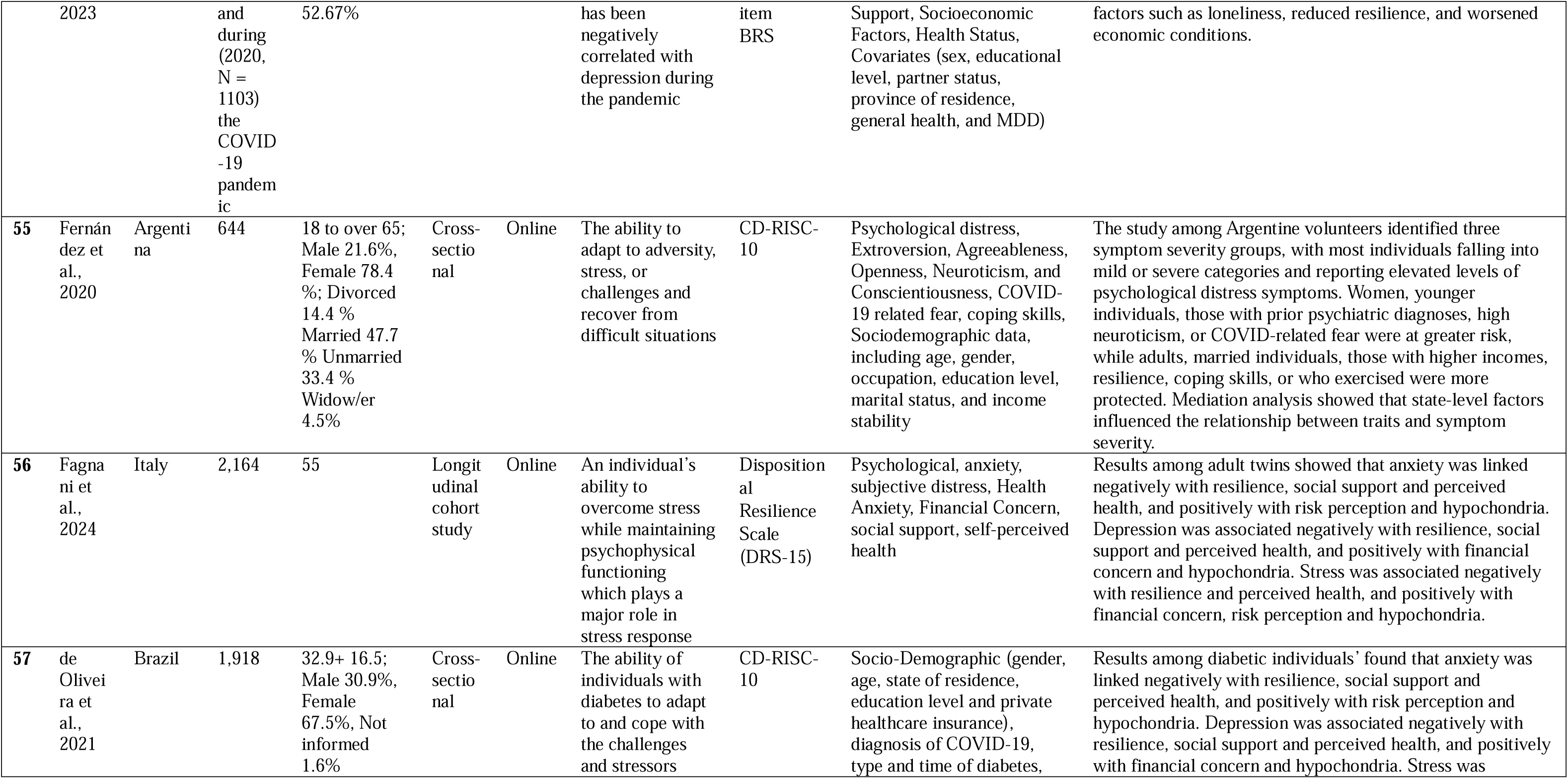
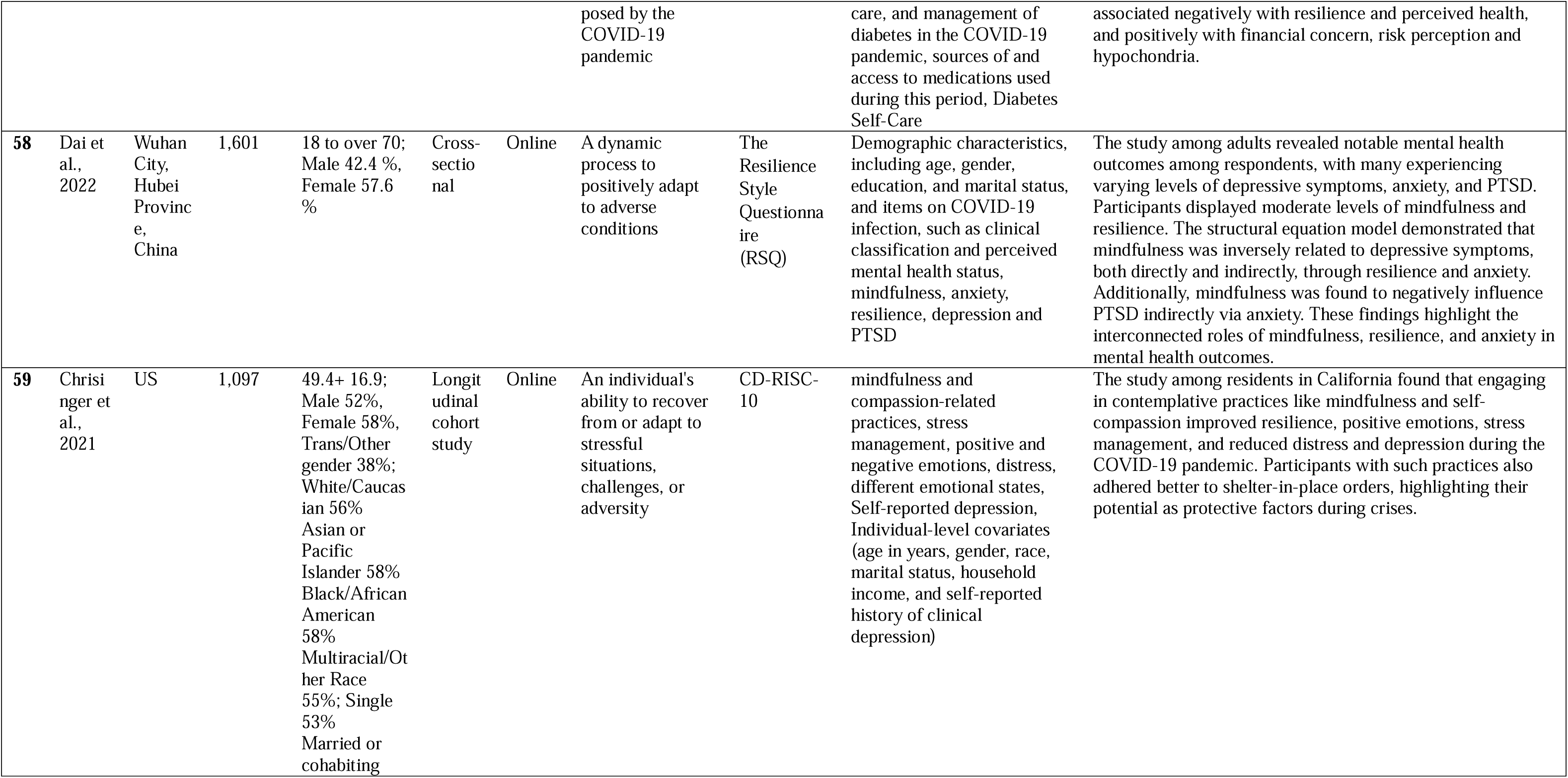
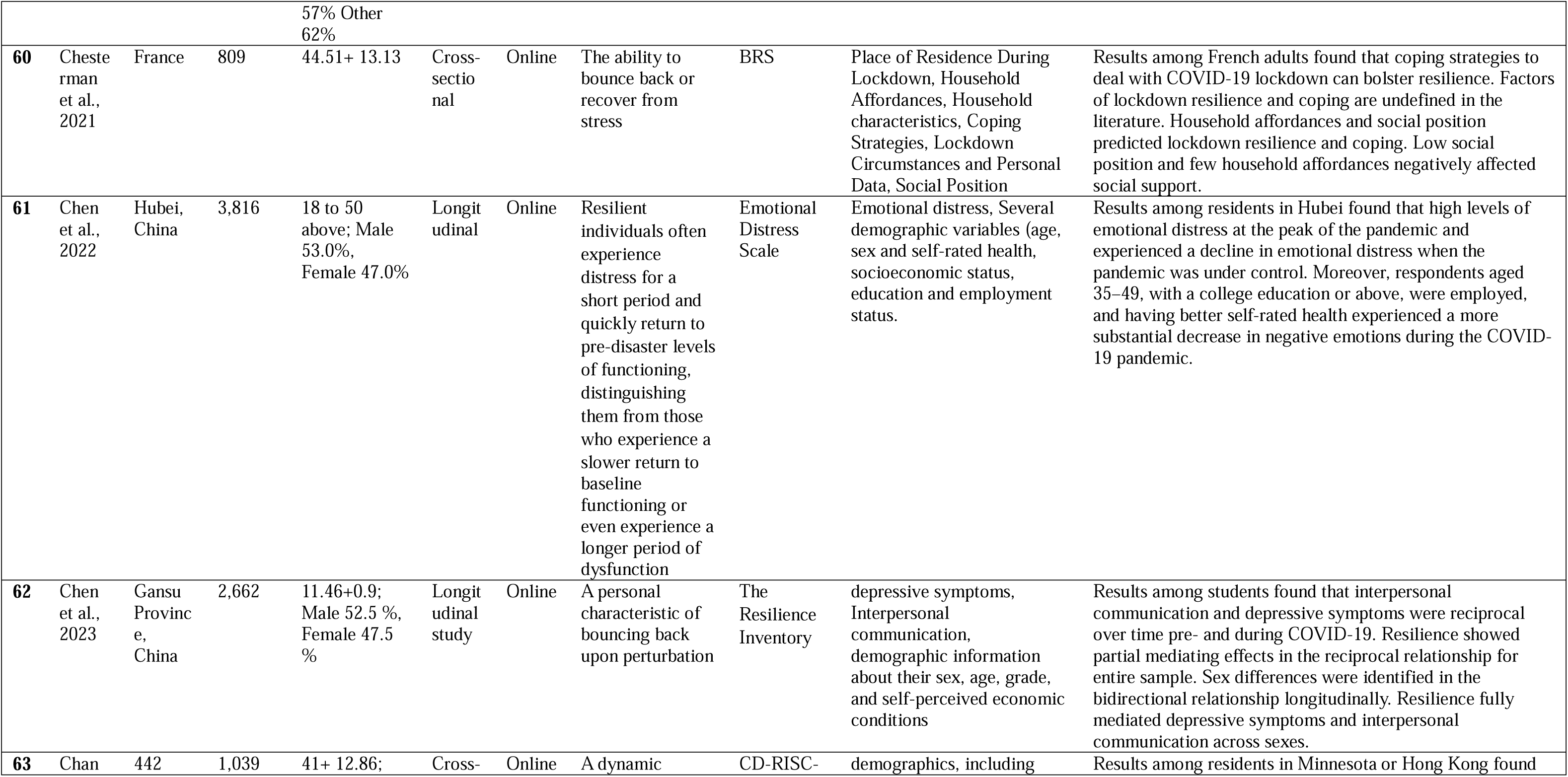
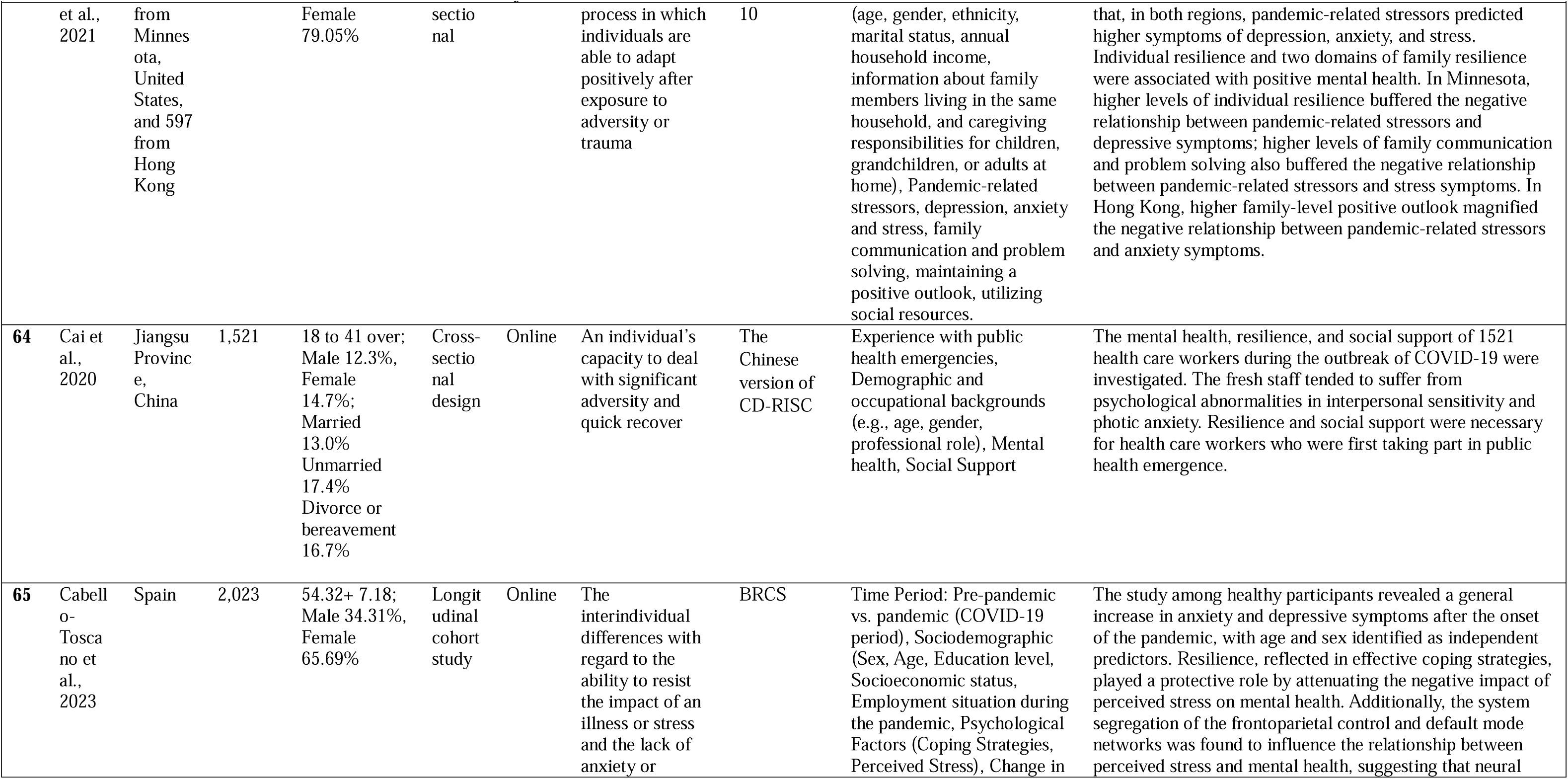
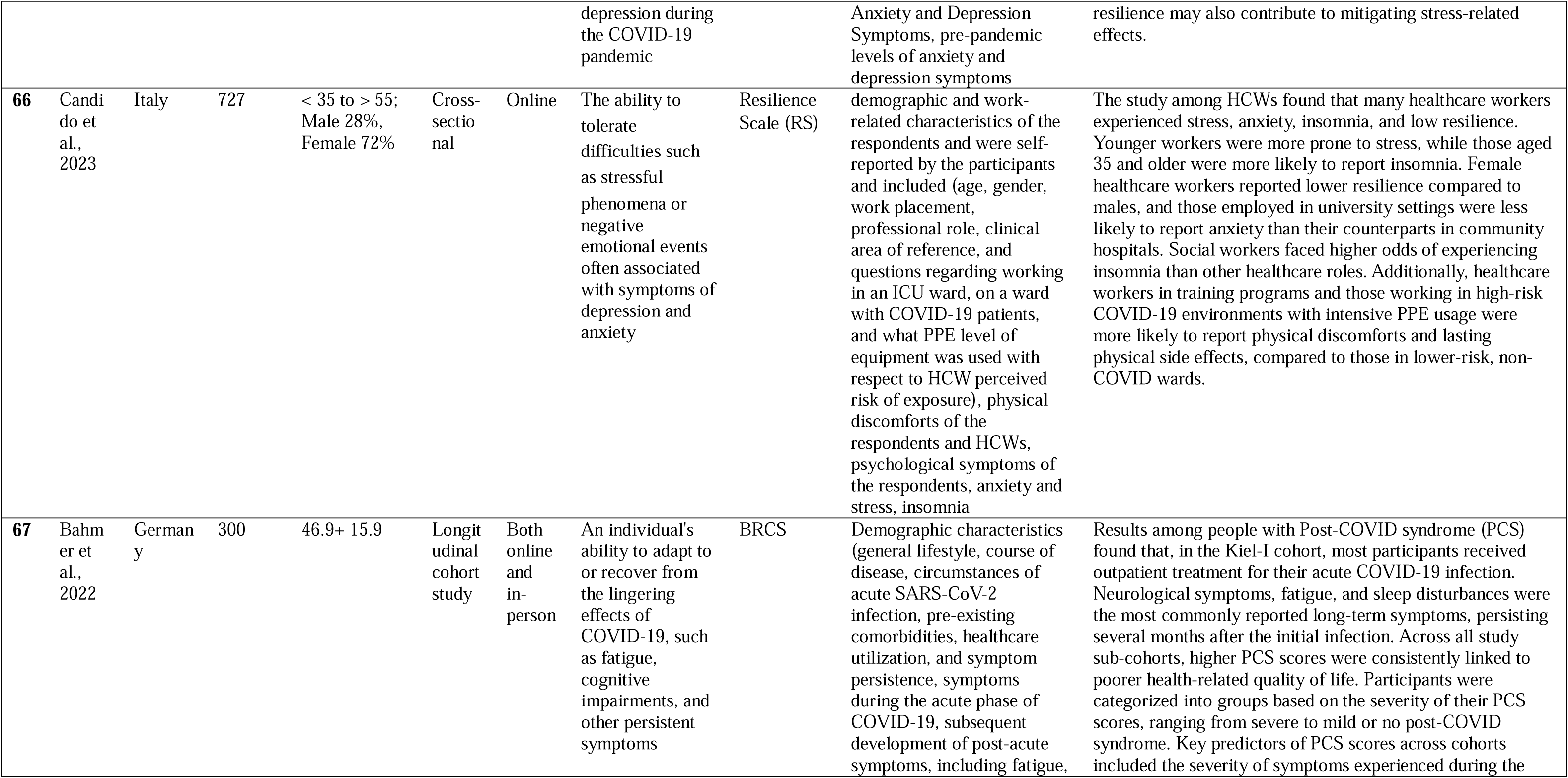
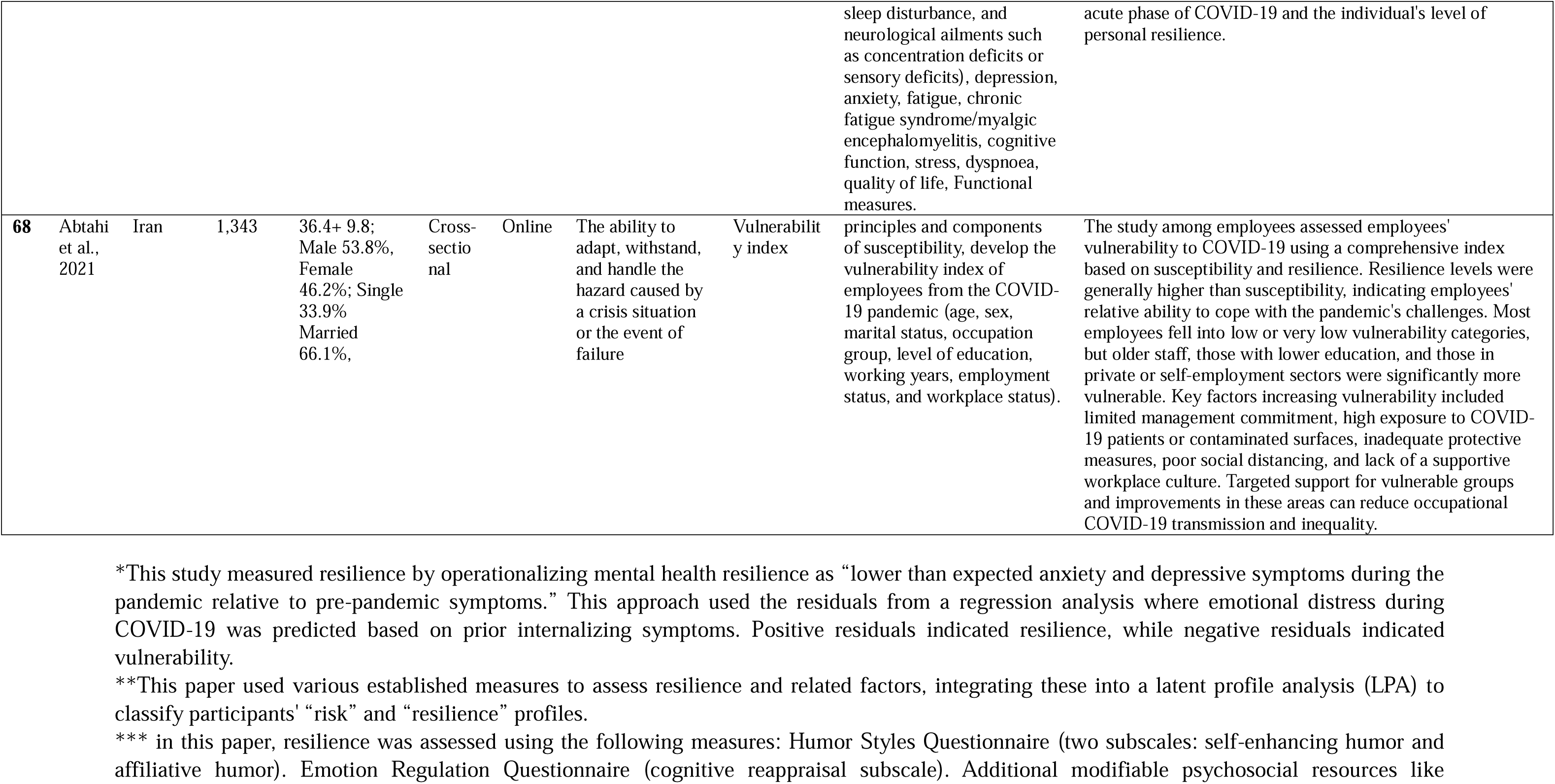
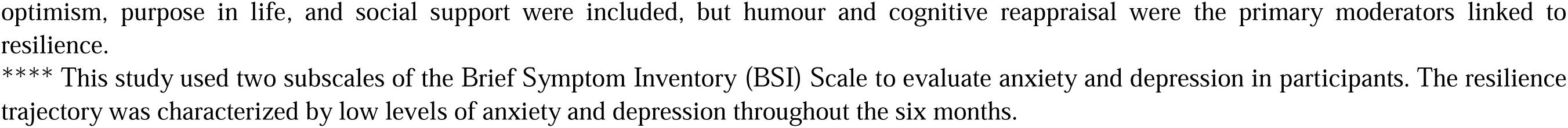
Summary of included studies.

The studies included in this review exhibited varying racial and ethnic compositions, which were influenced by the geographical location of the research. Studies conducted in Western countries like the United States, generally featured a more diverse sample, including a mix of Caucasian, African American, Asian, and Latino/Hispanic participants, reflecting the diverse demographics of the country. In contrast, studies conducted in Asia, such as in China, tended to feature a more homogeneous ethnic composition, predominantly consisting of the Han Chinese in China.

Age groups spanned from young people to older adults, with several studies specifically targeting specific demographic groups, which can be broadly categorized into essential workers during the pandemic and vulnerable populations. Essential workers were a significant focus, with studies examining healthcare workers (HCWs) (n=9) ^39–47^, school teachers and principals (n=4) ^48–51^ and quarantine hotel staff ^52^. Vulnerable populations included individuals with chronic diseases such as diabetes (n=1) ^53^, pregnant women (n=1) ^54^ and COVID-19 patients (n=4) ^55–58^. A considerable number of studies also examined younger populations, including college students (n=7) ^59–65^, elementary school students (n=2) ^66, 67^, high school students (n=2) ^61, 64^ and adolescents (n=3) ^64, 68, 69^. One study focused on twins (n=1) ^70^, presenting a unique perspective on organizational resilience that does not align neatly with the broader categories identified in this review. The included studies also featured a wide range of marital statuses and occupations, which further contributed to the diversity of the sample populations.

The sample sizes across the studies varied widely, ranging from a minimum of 61 participants ^71^ to a maximum of 14,269 participants ^72^. Most studies (n=49) employed a cross-sectional design, 16 utilized a longitudinal approach ^43, 55, 57, 61, 66, 70, 73–81^ and three adopted interventional designs ^59, 67, 69, 82^.

Most studies (n=61) employed online surveys as the primary data collection method. Three studies conducted data collection in person ^57, 58, 83^, while one used conducted phone calls ^84^, and three studies utilized a combined approach of online and in-person methods ^55, 67, 85^.

The included studies also featured a wide range of marital statuses and occupations, which further contributed to the diversity of the sample populations (Table 1).

### 3.2. Quality Assessment

Quality assessment results of various types of studies are summarized in Supplementary Tables 2a to 2c based on the study design (i.e., cross-sectional studies, longitudinal studies, and intervention studies).

#### 3.2.1 Cross-sectional studies

All 49 cross-sectional studies had clear research objectives (D1), participants from the same population (D4), clearly defined exposures (D9), and a clear definition of dependent variables (D11). Meanwhile, 98% (n=48) specified the study population (D2). Several studies failed to meet specific criteria, impacting their overall robustness and validity. Notably, 28 studies did not report related participation rates of eligible persons (D3), and 43 studies failed to state power analysis and sample size justification (D5). Also, nine studies did not explore various levels of exposure of interest in their studies (D8), and 18 studies did not control confounding variables (D14). Overall, as **Supplementary Table 2a** illustrates, 84% (n=41) of studies were rated as “Good” for having at least six “Yes” responses (out of nine) to the NIH tool dimensions. Eight were indicated as “Fair” for having 4-5 “Yes” responses, equaling 50% of all eligible dimensions.

#### 3.2.2. Longitudinal studies

Sixteen studies in this review were conducted using longitudinal study designs. All longitudinal studies could clearly state their research questions and objectives (D1) and their study population(D2) and had a sufficient timeframe for exploring the association between the exposures and the outcome (D7). They also clearly defined the exposures and used validated assessment measures (D10). However, 43% (n=7) of these studies failed to provide information regarding the participation rates (D3) and explored different levels of the interests’ exposures (D8). Meanwhile, 62.5% of them did not provide information about the sample size justification or power analysis. Overall, as **Supplementary Table 2b** demonstrates, 13 studies were rated as “Good” because of having at least nine “Yes,” and three studies were indicated as “Fair” if they had 6 to 8 “Yes,” which would equal at least 50% of all dimensions (based on eligible items).

#### 3.2.3. Interventional studies

This review included only three experimental studies: two randomized controlled trials (RCTs) and one pre-post test study without a control group. The quality assessment tool for the two RCTs was the Quality Assessment of Controlled Intervention Studies, while the tool used for the pre-posttest study was the Quality Assessment Tool for Before-After (Pre-Post) Studies With No Control Group.

RCT studies successfully addressed most of the NIH quality criteria, such as stating the study as RCT (D1), describing the method of randomization (D2), and keeping the overall dropout less than 20% (D7). On the other hand, these studies failed to provide information about whether participants were blind to treatment group assignments (D4), as stated if the outcome assessors were also blinded to group assignments of the participants (D5), and they did not use intention-to-treat analysis (D14). However, both studies were rated as “Good” as they addressed nine of the NIH tool criteria, as shown in **Supplementary Table 2c**. Our last study was also rated as “Good,” as 63% of NIH criteria were addressed and received “Yes” to the relevant dimensions. The methodology was strong regarding a clear study aim (D1), participants who were suitable representatives of the general populations (D2), assessing outcomes changing before the interventions to after them (D10), and other dimensions, as shown in **Supplementary Table 2c**.

### 3.3. Resilience Definition and Measurement

Psychological resilience has been defined in various ways across the literature. It is broadly characterized as the ability to positively adapt to stress, adversity, or crises^40, 42, 45, 46, 51, 52, 59, 62, 63, 65, 67, 69-72, 77, 78, 82, 83, 85-94^ and to cope effectively with challenging situations ^42, 63^. Resilience refers to maintaining both physical and mental health during times of heightened stress ^48, 95, 96^, functioning as a protective factor in such contexts ^43, 50, 58, 60, 61, 86, 91, 96^. Some studies review resilience as a process, a multifaceted construct, and a dynamic process influenced by an interplay of biological, social, cultural, and psychological factors ^56, 60, 76, 97^. This complexity reflects its evolving nature and highlights its significance in both individual and collective responses to adversity.

A wide range of scales was employed across the studies to measure resilience, with the Connor–Davidson Resilience Scale (CD-RISC) and its various versions being the most commonly used tool (n=28), including adapted versions like the Chinese and Arabic CD-RISC ^39, 40, 43–45, 47, 50, 52–54, 58–61, 64, 65, 67, 69, 81–84, 89, 97–101^, with a Cronbach’s alpha values of between 0.76 and 0.97. The Brief Resilience Scale (BRS) was utilized by 15 studies^51, 55, 62, 63, 71, 72, 74, 77–79, 87, 88, 90, 96, 102^, with Cronbach’s alpha ranging from 0.70 to 0.92. The Resilience Scale for Adults (RSA) appeared in three studies ^57, 73, 103^ with Cronbach’s alpha ranging from 0.67 to 0.90. Other used measures included the Brief Resilient Coping Scale (BRCS) (n=2) ^95, 104^, with Cronbach’s alpha of 0.65 to 0.69, and the Ego-Resiliency Scale (ER89) (n=2)^94, 105^, with a Cronbach’s alpha of 0.78 to 0.90. Additionally, several studies (n=1 each) employed a variety of other scales, such as the Resilience Evaluation Scale (RES)^46^ with Cronbach’s alpha of 0.825^106^, and the Dispositional Resilience Scale (DRS-15)^70^, with Cronbach’s alpha 0.73^107^.

### 3.4. Factors Associated with Psychological Resilience

This review identified factors that may influence resilience, which were grouped into demographic, individual, interpersonal, community/ organizational, and structural levels (See **Table 1**).

#### 3.4.1. Demographics

Younger individuals tend to exhibit lower resilience, ^74, 78, 88, 100^. Research indicates that younger adults, due to their higher susceptibility to depression, tend to exhibit lower levels of resilience, with depression undermining the development of resilience and mediating the relationship between age and resilience^74^. Younger adults experience greater vulnerability to stress and its psychological effects, possibly due to limited experience in managing adversity^74^. In contrast, studies have shown that older adults, particularly those aged 60 or above, experienced fewer pandemic-related stressors than their younger counterparts, potentially due to factors like retirement, which made social distancing less intrusive. These reduced stressors and life circumstances likely contributed to higher levels of resilience among older adults, enabling them to cope more effectively with pandemic-related challenges ^97^. These findings suggest that resilience increases with age probably as a result of reduced exposure to stressors and life experience, which enhances their ability to mitigate mental health difficulties.

Resilience also exhibits gender-related differences, although findings across studies have been mixed. Some studies have suggested that women demonstrate greater resilience compared to men, possibly influenced by stronger social support networks that foster resilience during adversity^66^. However, other research points to contrasting results, where women report lower resilience than men. For instance, a study focusing on HCWs found that female HCWs exhibited lower resilience compared to their male counterparts^41^, suggesting that the demands and stressors within certain professions might negatively impact the development or maintenance of resilience. It is also possible that more nurses being female may be exposing them to a larger workload of directly taking care of patients, which could contribute to these observed gender differences in resilience among HCWs.

Certain groups, including HCWs and teachers, experienced unique stressors that further hindered resilience. High work demands and physical strain led to burnout and exhaustion ^108^. For example, oncology professionals reported increased work hours and job insecurity, which heightened emotional exhaustion and reduced resilience ^109^. Similarly, nurses in mobile cabin hospitals faced unfamiliar environments, heavy workloads, and elevated infection risks, resulting in severe psychological trauma and lower resilience levels ^45^. Teachers also faced significant challenges during COVID-19, particularly related to work-family conflicts stemming from the rapid transition to remote teaching. The shift to online teaching blurred the boundaries between personal and professional life, leading to increased emotional exhaustion and stress, which negatively impacted resilience outcomes. For instance, Manuti et al. found that the work-life interface became increasingly problematic as teachers struggled to balance their responsibilities as both educators and parents. This transition required teachers to adapt quickly to new teaching environments, often from home, leading to heightened work-family conflicts and strained their mental health ^49^.

#### 3.4.2. Individual level

Resilience during the COVID-19 pandemic was shaped by a complex interplay of psychological, personality, and behavioral factors, which could either facilitate or impede psychological resilience. Mental health conditions such as anxiety, depression, and burnout were consistently associated with lower resilience levels, as they depleted cognitive resources necessary for adaptive coping ^41, 42, 50, 63, 64, 69, 71, 76, 78,79, 92, 94, 98, 99, 102, 105^. These mental health challenges were particularly evident among caregivers, parents, HCWs, students, and teachers who struggled to balance caregiving, remote learning, and work responsibilities amidst heightened health and financial concerns. Vaccine uptake and substance use also influenced resilience. Vaccinated individuals reported greater resilience outcomes, likely due to reduced anxieties about infection and increased perceptions of safety ^47^. Conversely, substance use was negatively associated with resilience, as individuals who engaged in substance use were less likely to develop higher resilience levels ^42, 55, 83, 104^.

Despite these challenges, several factors were identified as protective influences on resilience. Adaptive personality traits, such as emotional regulation, self-efficacy, and optimism, were strongly associated with improved resilience outcomes during the pandemic ^47, 57, 63, 67, 69, 71, 87, 89, 91, 96, 100^. Individuals who employed effective coping mechanisms, such as self-regulation, acceptance, self-compassion, and exercise, demonstrated greater resilience levels, buffering the negative effects of stress and anxiety ^42, 55, 83, 104^. Conversely, negative thought patterns, such as rumination on health risks and uncertainty about the future, amplified feelings of helplessness and were linked to lower resilience levels ^62, 72, 103^. Interestingly, travel-related fears during the pandemic prompted some individuals to develop coping strategies, which further contributed to improved resilience levels by helping them manage emotional distress ^89^.

Physical activity played a particularly significant role in fostering resilience. Regular exercise and home-based routines were consistently associated with higher resilience levels and reduced symptoms of depression and anxiety. For instance, engaging in physical activity enhanced resilience among young adults and helped individuals manage stress more effectively during the pandemic ^96, 100^.

#### 3.4.3. Interpersonal level

Resilience during the COVID-19 pandemic was profoundly influenced by interpersonal factors, including the availability of social support, the quality of interpersonal relationships, and the impacts of social isolation and work-life conflicts. The pandemic’s restrictions, including lockdowns and social distancing measures, exacerbated loneliness and isolation, further hindering resilience. Studies highlighted that reduced opportunities for in-person interactions limited individuals’ ability to rely on external social resources, leading to feelings of isolation and diminished psychological resilience outcomes^40, 45, 51, 64, 65, 71,78, 81, 88, 105^.

Social support emerged as a critical factor in fostering resilience. Strong interpersonal relationships and engagement in social activities significantly enhanced individuals’ coping abilities and psychological resilience ^40, 44, 48, 57, 61, 63–65, 87, 99, 104, 105^. For instance, during the first quarantine in France in 2020, Rotonda et al. (2021) found that daily social contact, facilitated by increased use of video calls and messaging, helped sustain psychological well-being and foster higher resilience ^87^. Similarly, a study on Chinese adolescents showed that greater levels of perceived social support were associated with improved resilience during the lockdown ^64^.

The absence of structured psychological and social support systems negatively impacted resilience, with 81% of respondents in one study reporting that they did not receive needed support, even though 88% recognized its importance ^41^. Families with members who had special educational needs and disabilities (SEND) faced heightened stress due to the loss of access to typical support systems, such as external caregivers and professional services, which contributed to lower resilience levels ^71^.

#### 3.4.4. Community and organizational level

Resilience during the COVID-19 pandemic was significantly shaped by community and institutional factors, including the availability of social support within institutions, workplace conditions, and institutional responses to the crisis. The suspension of community religious activities, particularly communal prayers, removed a vital source of emotional and social support for many individuals, particularly the elderly. For example, in Muslim-majority countries like Qatar, the closure of places of worship disrupted religiosity as a crucial coping mechanism, leading to heightened levels of anxiety and depression among elderly populations. This lack of community-based spiritual support negatively influenced resilience outcomes for vulnerable groups, emphasizing the essential role of communal and spiritual coping strategies in promoting psychological resilience during adversity ^84^.

Workplace environments, particularly for HCWs, played a central role in shaping resilience outcomes ^108^. High workloads, inadequate personal protective equipment (PPE), and constantly evolving safety protocols created significant challenges for healthcare professionals ^41–44, 46–48, 97^. For instance, Italian HCWs in high-risk COVID-19 wards reported severe stress, anxiety, and insomnia, largely attributed to physically demanding PPE requirements and overwhelming responsibilities ^41^. Similarly, Brazilian physiotherapists working with COVID-19 patients experienced elevated levels of emotional exhaustion, depression, and anxiety due to long shifts, inadequate PPE, and the constant fear of exposure. These stressors strongly correlated with diminished resilience outcomes among HCWs ^42^.

However, workplaces that prioritized employee health through effective top management practices helped mitigate these impacts. Providing essential resources, such as PPE, handwashing facilities, and clear guidance on COVID-19 risks, significantly enhanced resilience by alleviating workplace stress and instilling confidence in organizational responses ^85^. It seems that workplaces that ensured job satisfaction and vaccination availability among HCWs contributed to greater resilience, both for employees and the communities they served ^47^.

#### 3.4.5. Structural level

Structural factors, such as public health resources, economic conditions, and accessibility to mental health services, played a critical role in shaping resilience outcomes during the COVID-19 pandemic. Limitations in public health infrastructure, including a shortage of trained mental health professionals and inadequate resources, hindered the widespread delivery of resilience interventions, particularly in rural and underserved regions ^41, 57, 58, 60, 66, 70, 74, 77, 80, 93, 94^. For instance, public health systems in Spain struggled to meet psychological care demands, with insufficient professional-to-patient ratios and overwhelmed services negatively impacting the quality and accessibility of mental health care ^74^. These systemic barriers significantly constrained individuals’ ability to access support, contributing to diminished resilience levels among vulnerable populations.

Economic hardships exacerbated these structural challenges, further compounding emotional distress and reducing individuals’ capacity to foster resilience. Socioeconomic status emerged as a critical determinant of resilience outcomes during the pandemic. Individuals with higher socioeconomic status demonstrated greater resilience outcomes, benefiting from increased access to stress management resources and mental health support. Stable incomes and belonging to an upper-income class were associated with enhanced coping capacities and improved resilience levels, enabling individuals to better cope with the challenges of the pandemic ^100^. In contrast, those in lower socioeconomic classes faced significant barriers, including financial insecurity and limited access to resources, which were strongly linked to diminished resilience outcomes ^90^.

Financial insecurity, worsened by job losses, precarious incomes, and lockdown measures, disproportionately affected low-income and single-parent families. Restricted work opportunities during the pandemic intensified psychological stress, leading to reduced compliance with protective measures and lower resilience outcomes^52, 61, 87, 90, 98, 101^. A study in Argentina highlighted that individuals with inconsistent incomes were more likely to experience elevated psychological distress, whereas those with stable incomes and higher socioeconomic status demonstrated greater resilience levels and enhanced coping capacities, mitigating the adverse effects of pandemic-related stressors ^100^. Although government support programs provided financial relief for vulnerable households, these interventions often failed to address the emotional distress associated with financial insecurity^93^.

Finally, stigma and mistrust in mental health services posed additional barriers to resilience. A study in New York on mental health screening for older adults revealed that only 14% of referrals were accepted, while 86% were declined, with most rejections attributed to outright refusal of services. For individuals such as people with disabilities (PWDs) and the elderly, limited access to essential healthcare and rehabilitation services, compounded by pandemic restrictions, further diminished resilience outcomes. Social distancing measures disrupted caregiving routines and access to necessary daily support, leaving these populations more vulnerable to psychological distress ^77, 91^.

### 3.5. Interventions to Improve Psychological Resilience and Their Efficacy

As shown in **Table 2**, only three out of the 68 included studies focused on interventions to enhance resilience during the COVID-19 pandemic. Two of these studies evaluated the efficacy of mindfulness and emotional intelligence training (n=180) ^69^ and the Positive Interpersonal and Life Orientation Training (PILOT) program (n=868)^67^.The third study assessed the impact of media exposure on resilience and emotional states during the pandemic (n=175) ^59^. Two studies targeted young populations, with one focusing on adolescents (n=180)^69^ and the other on elementary school students (n=868)^67^. The third study focused on the general adult population^59^ (n=175). Key intervention components across these studies included emotional regulation, cognitive skills, and systemic support.

**Table 2.**
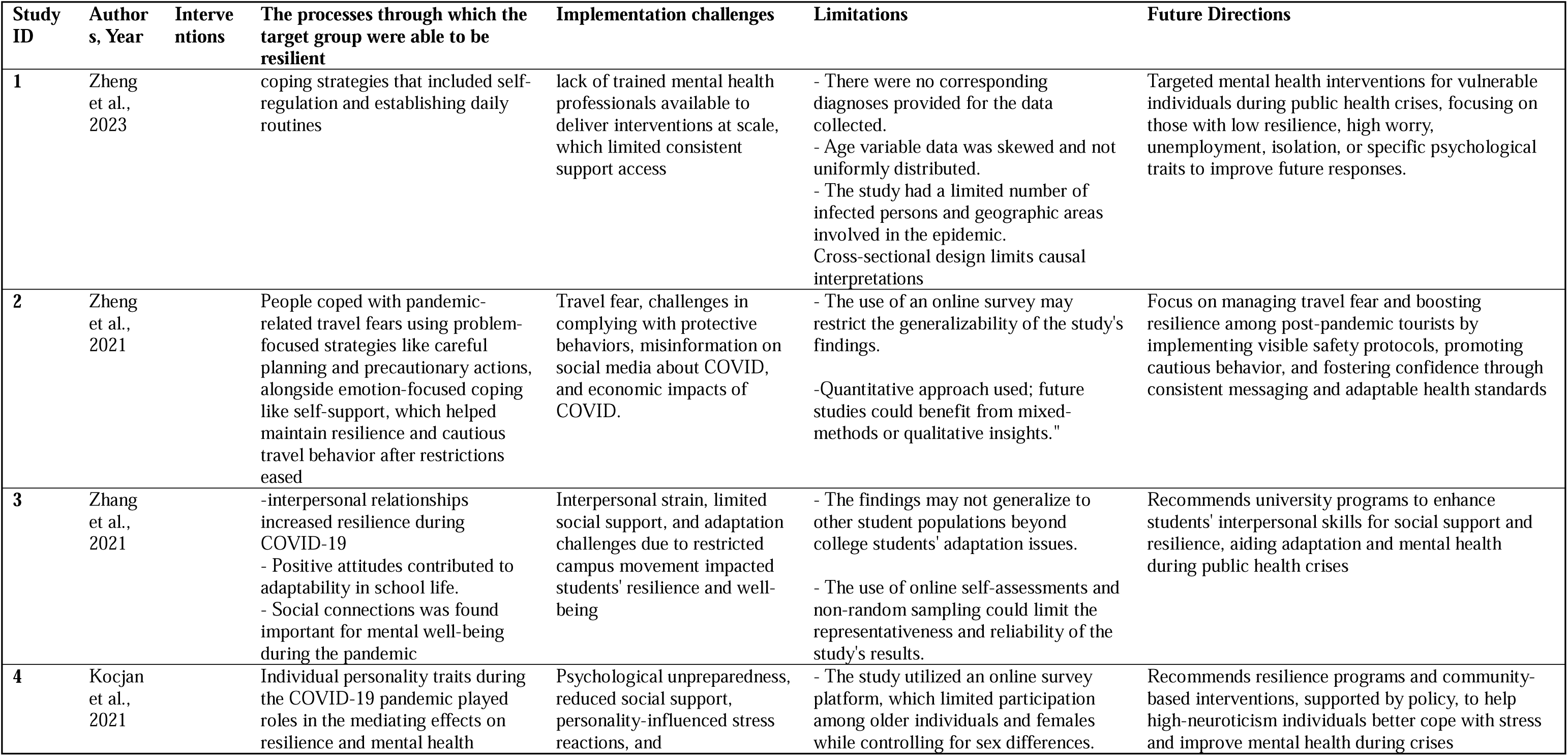
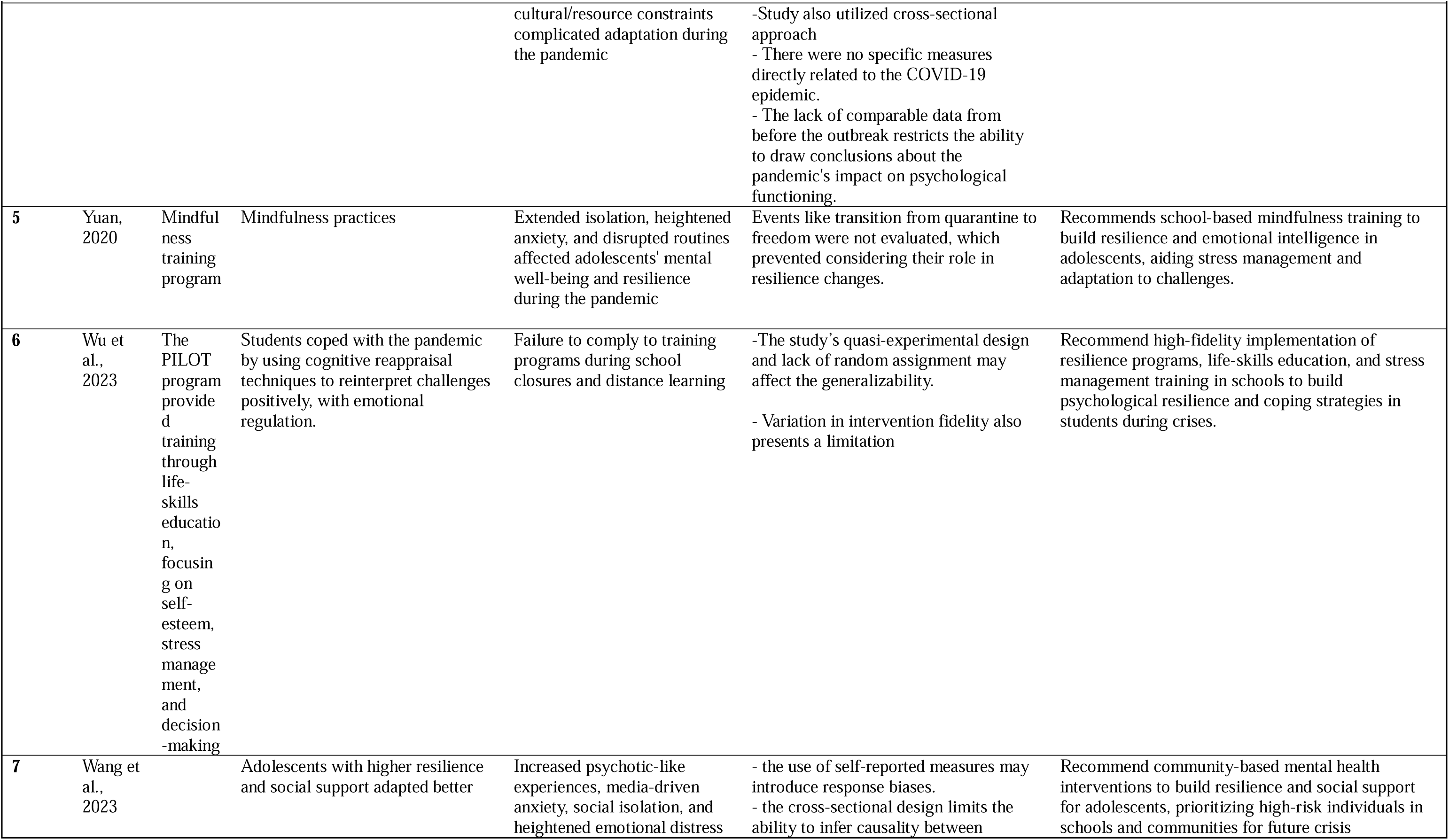
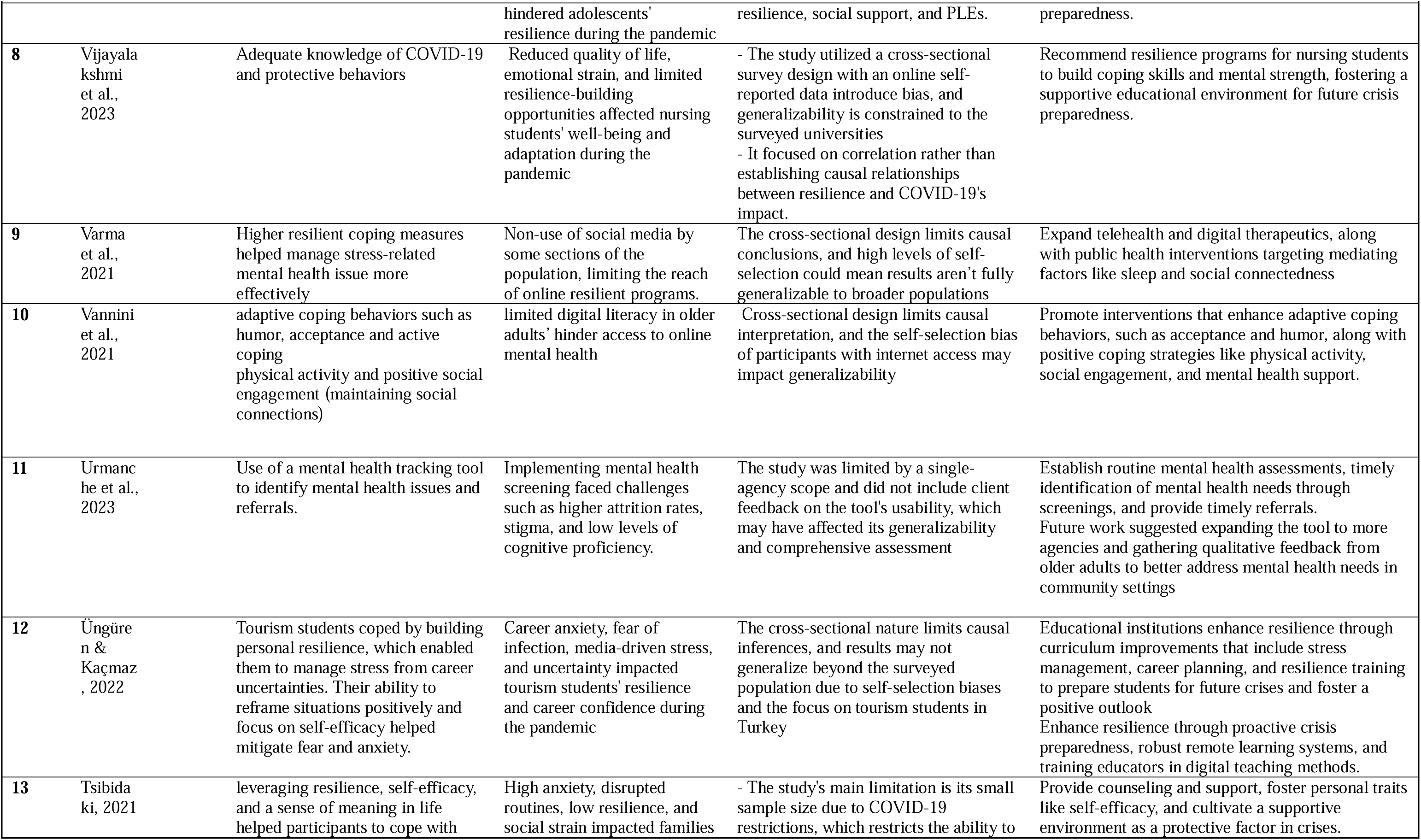
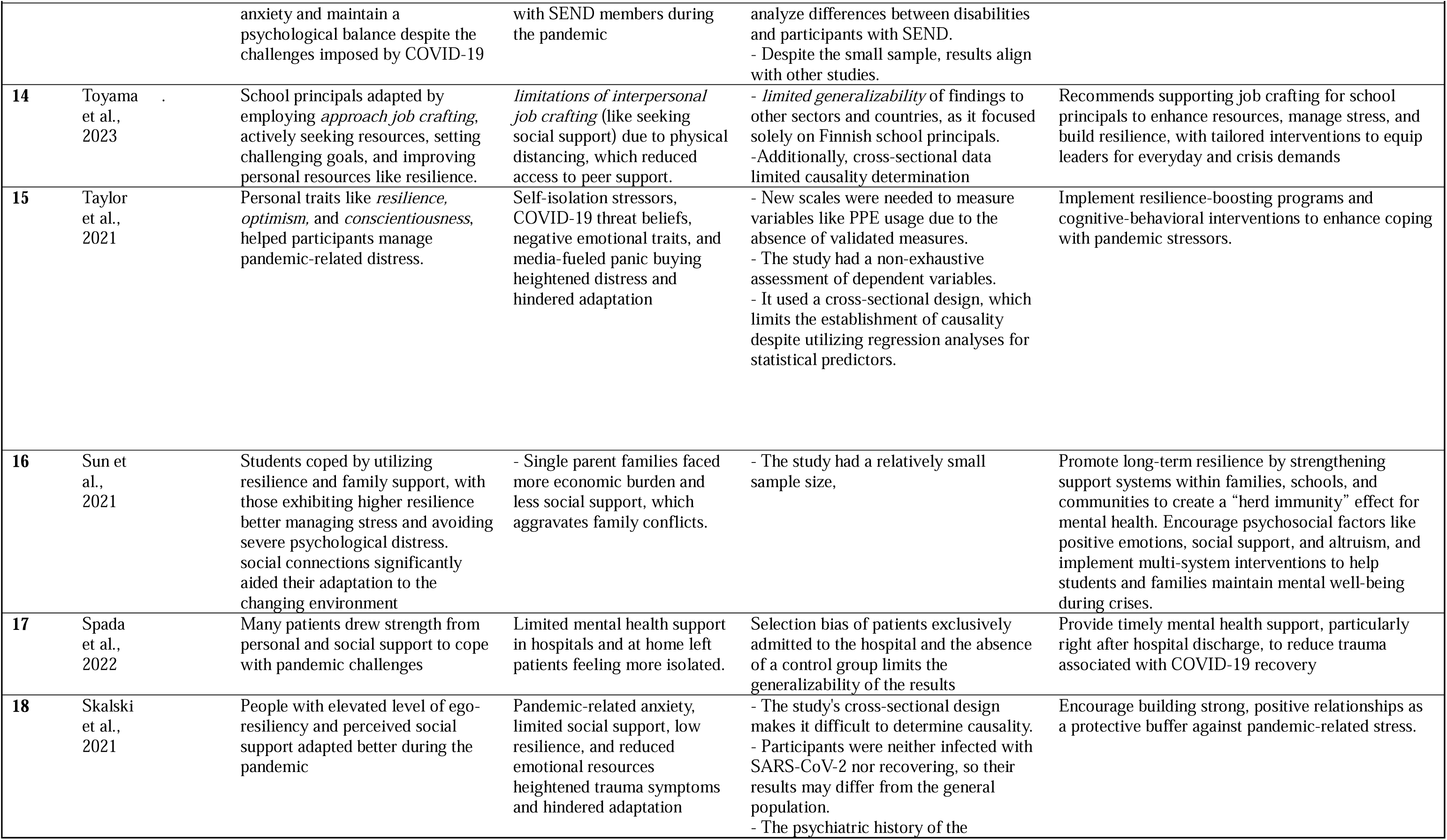
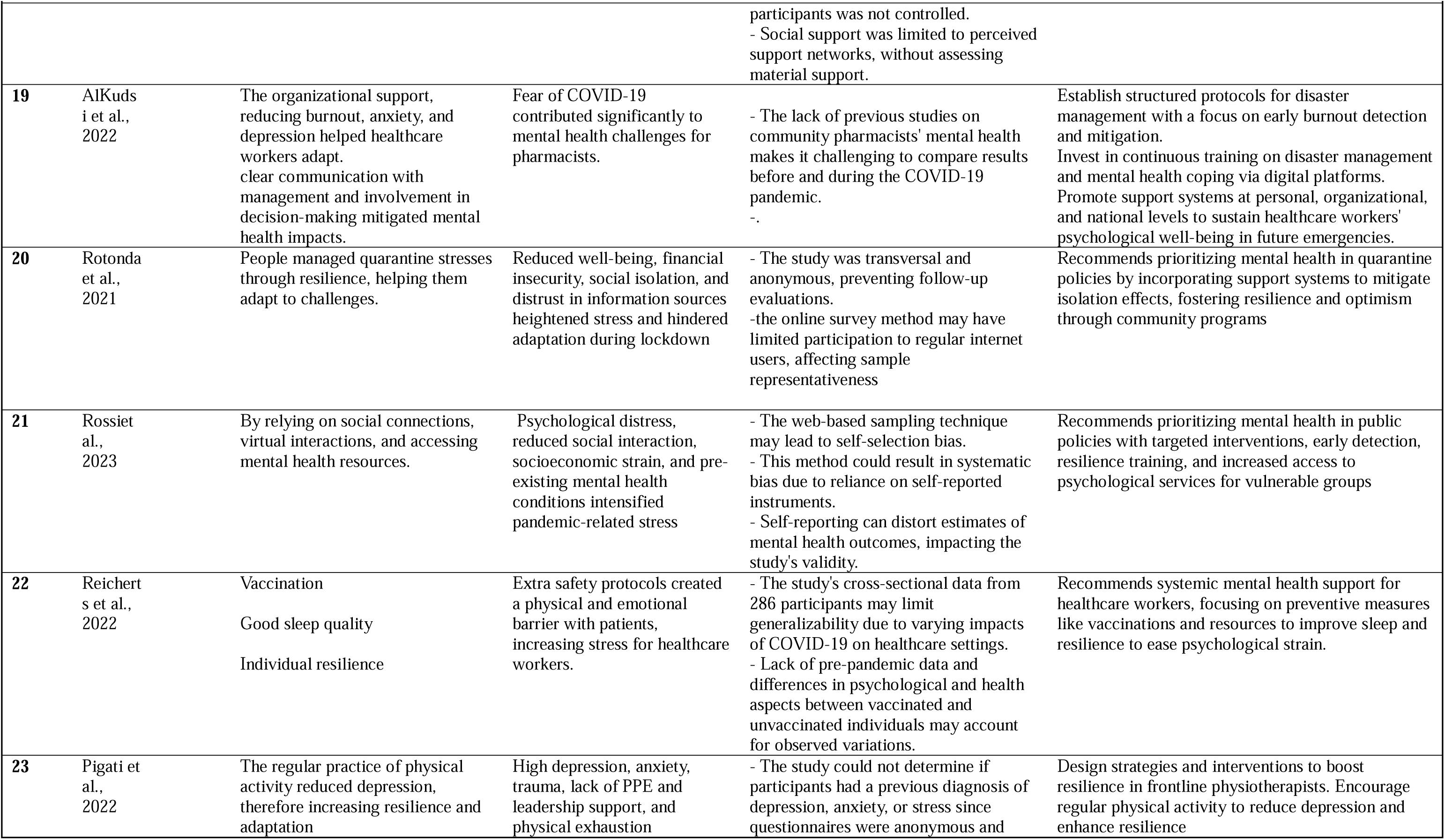
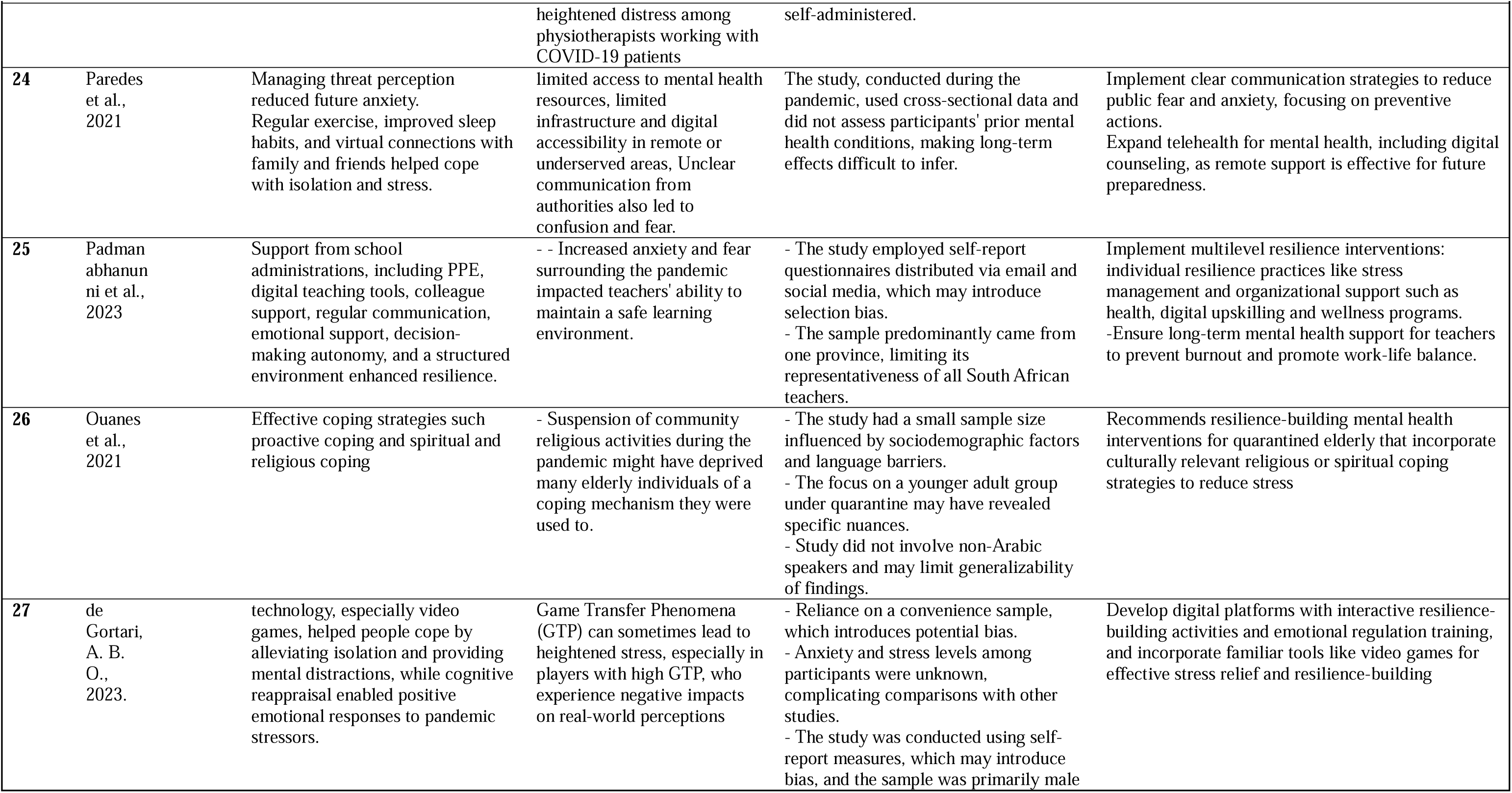
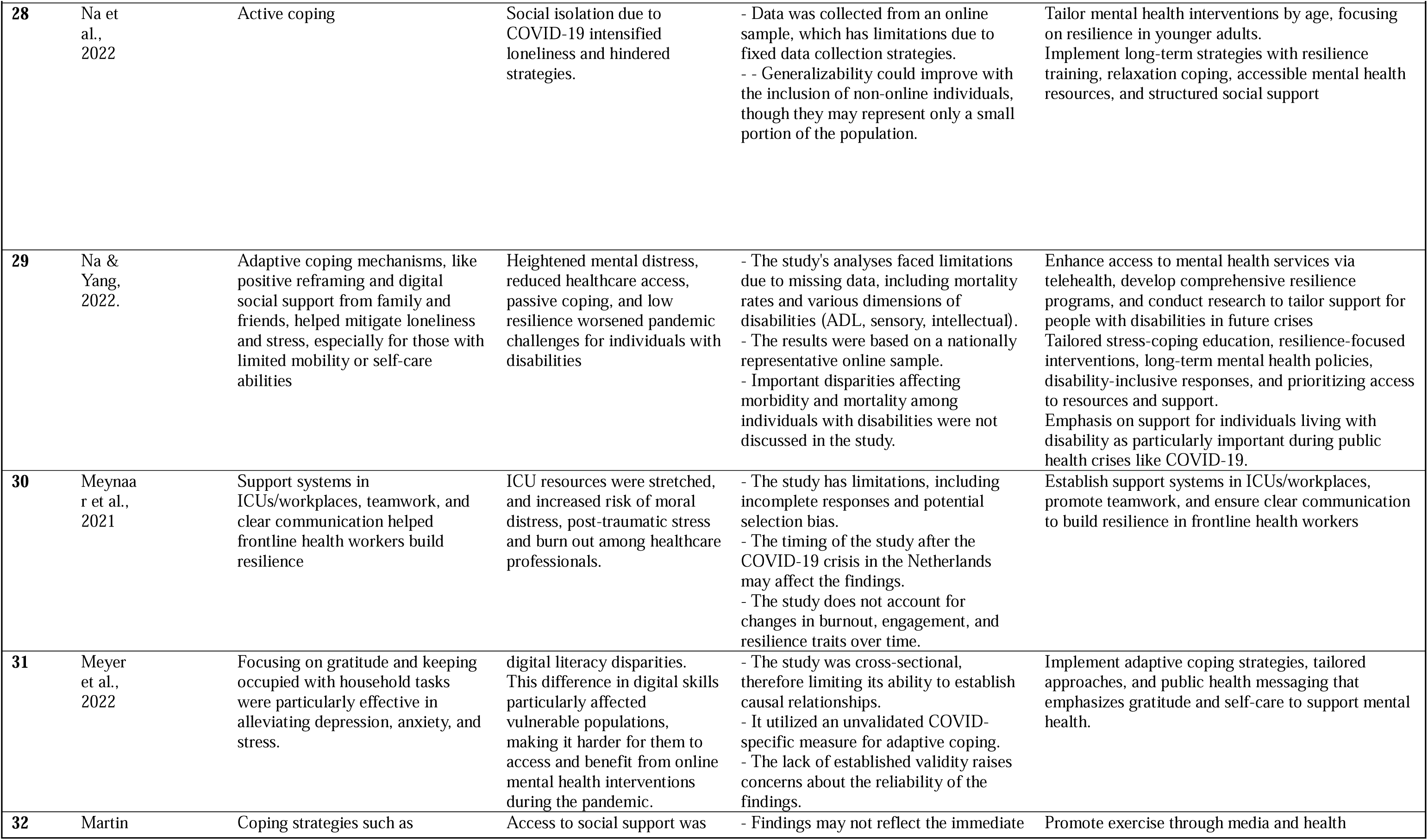
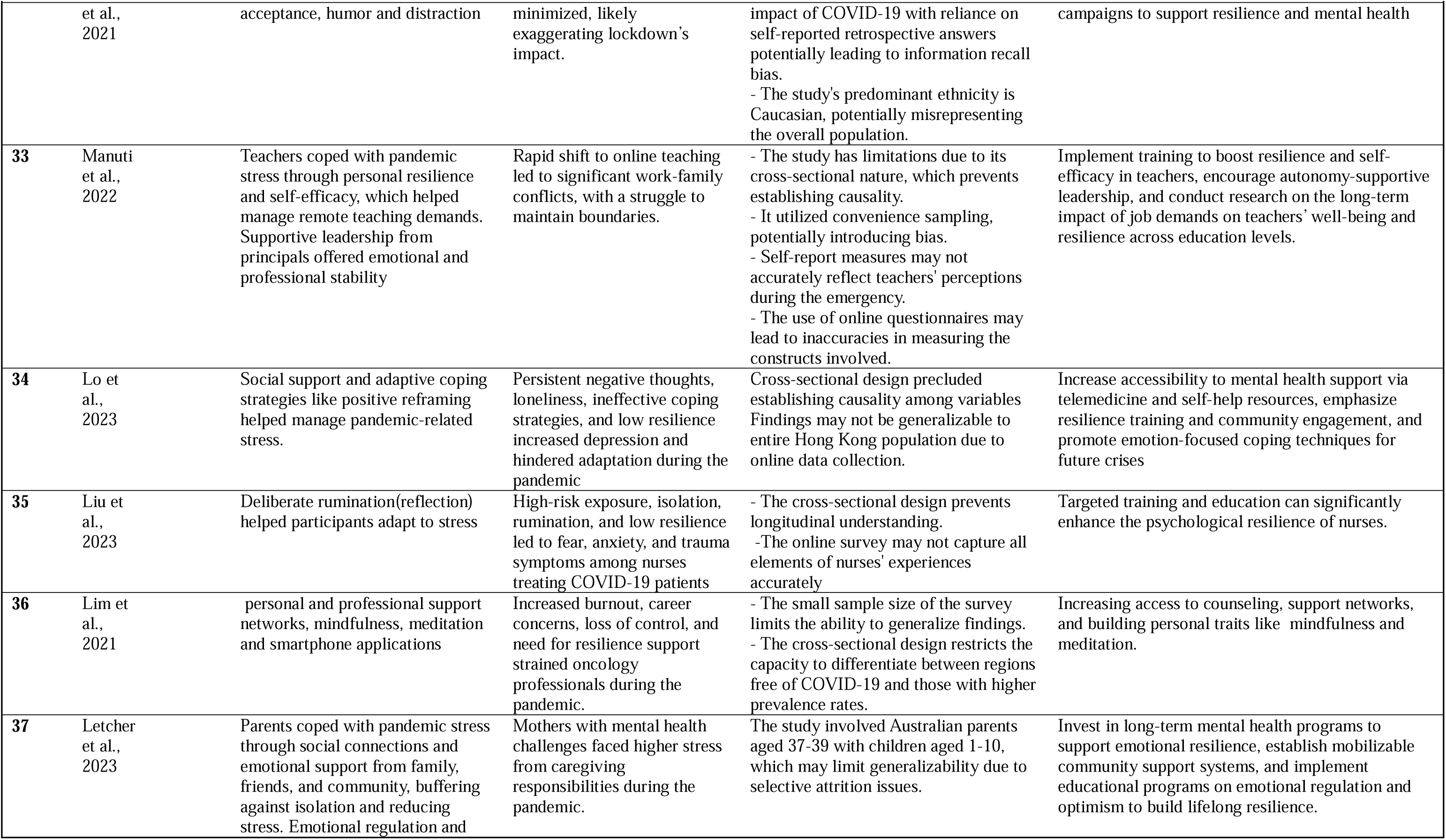
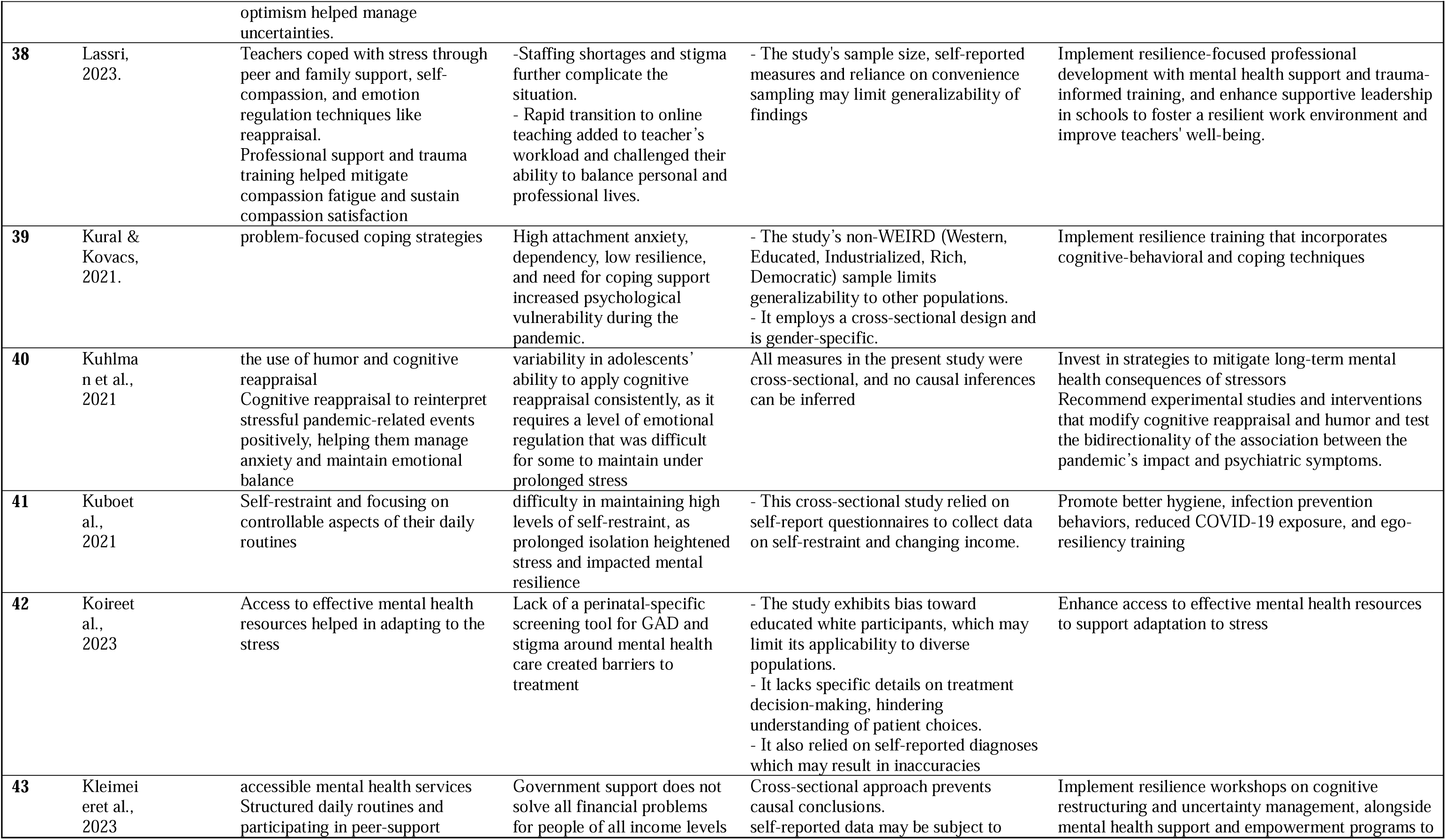
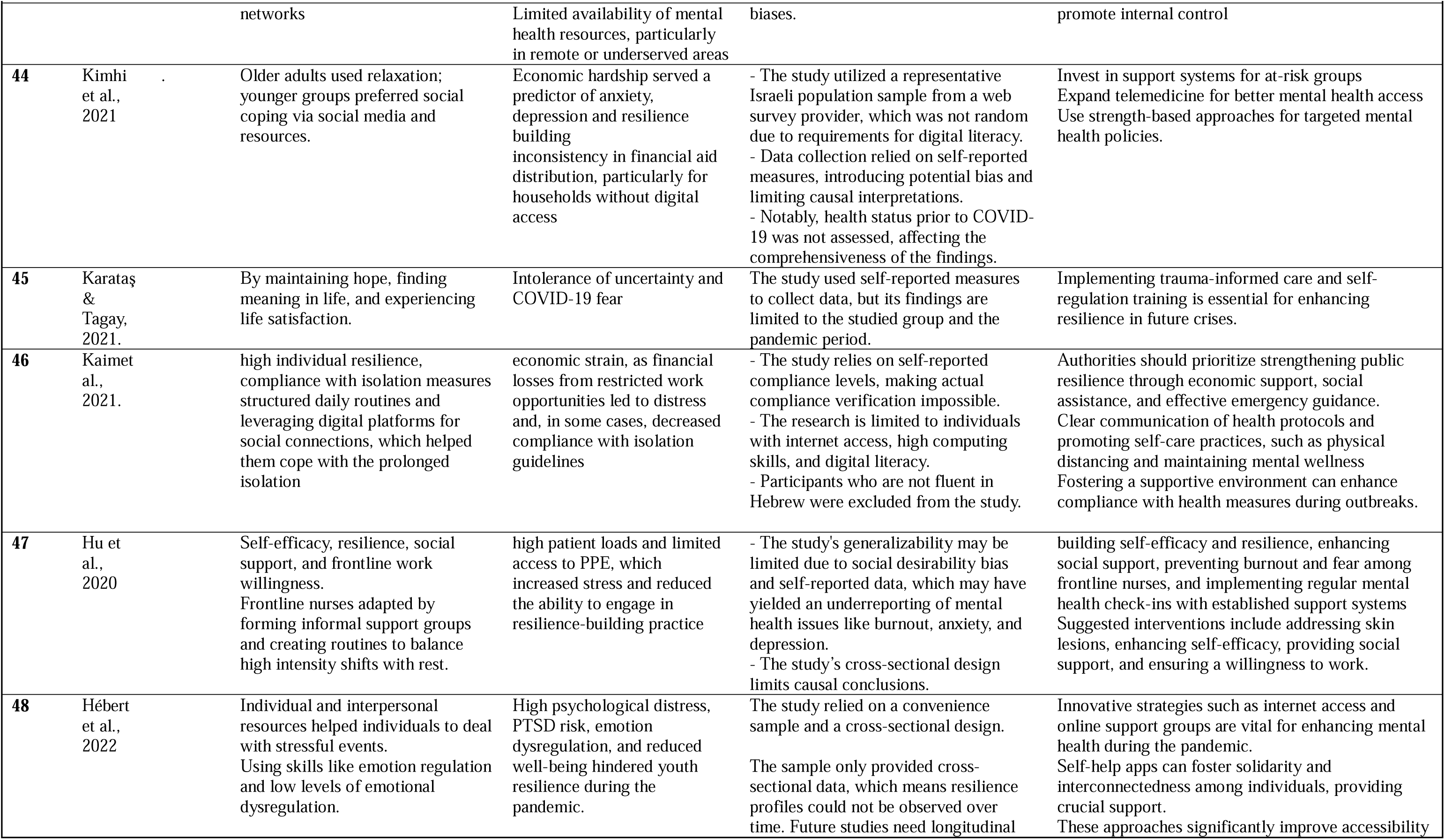
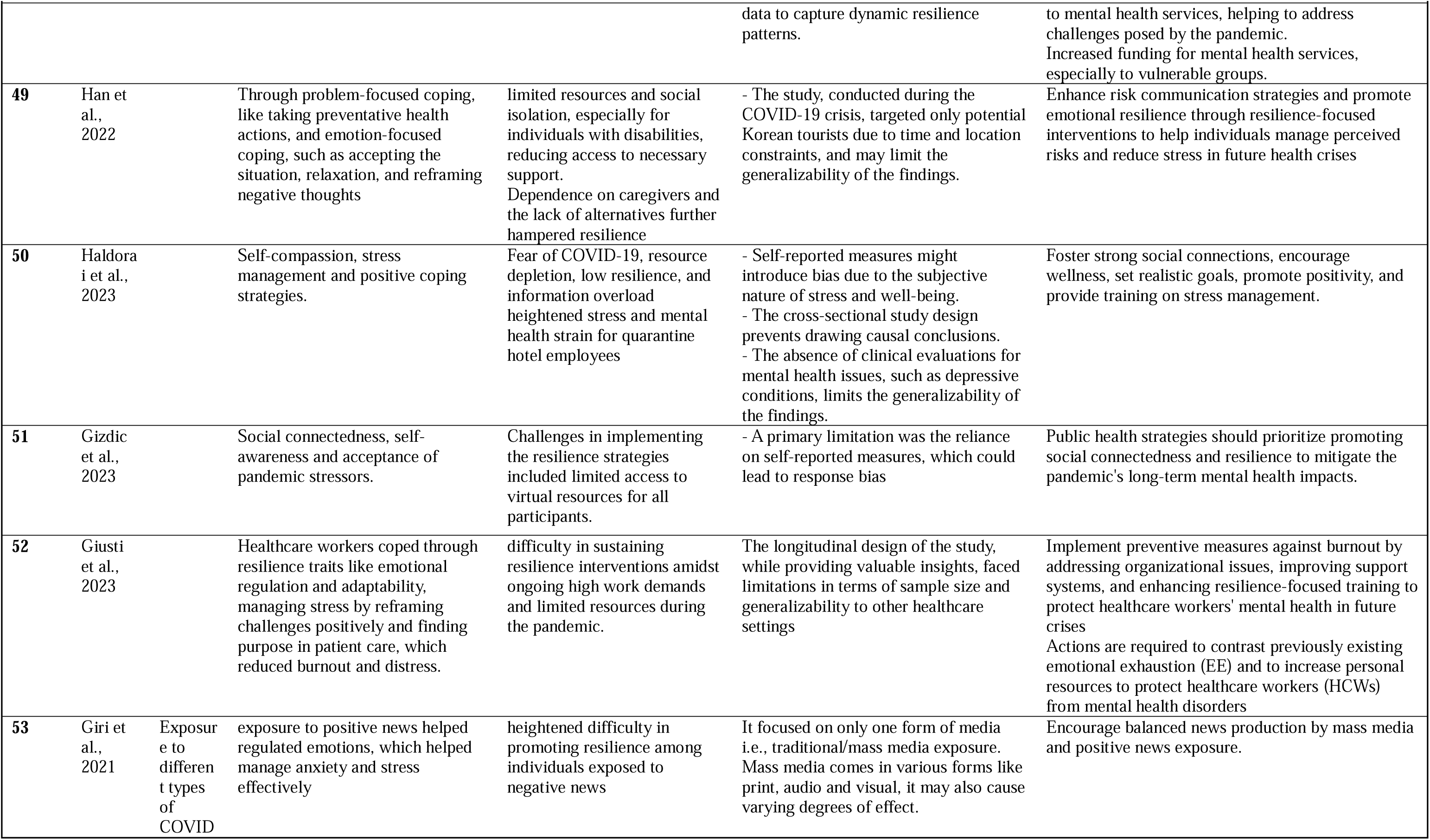
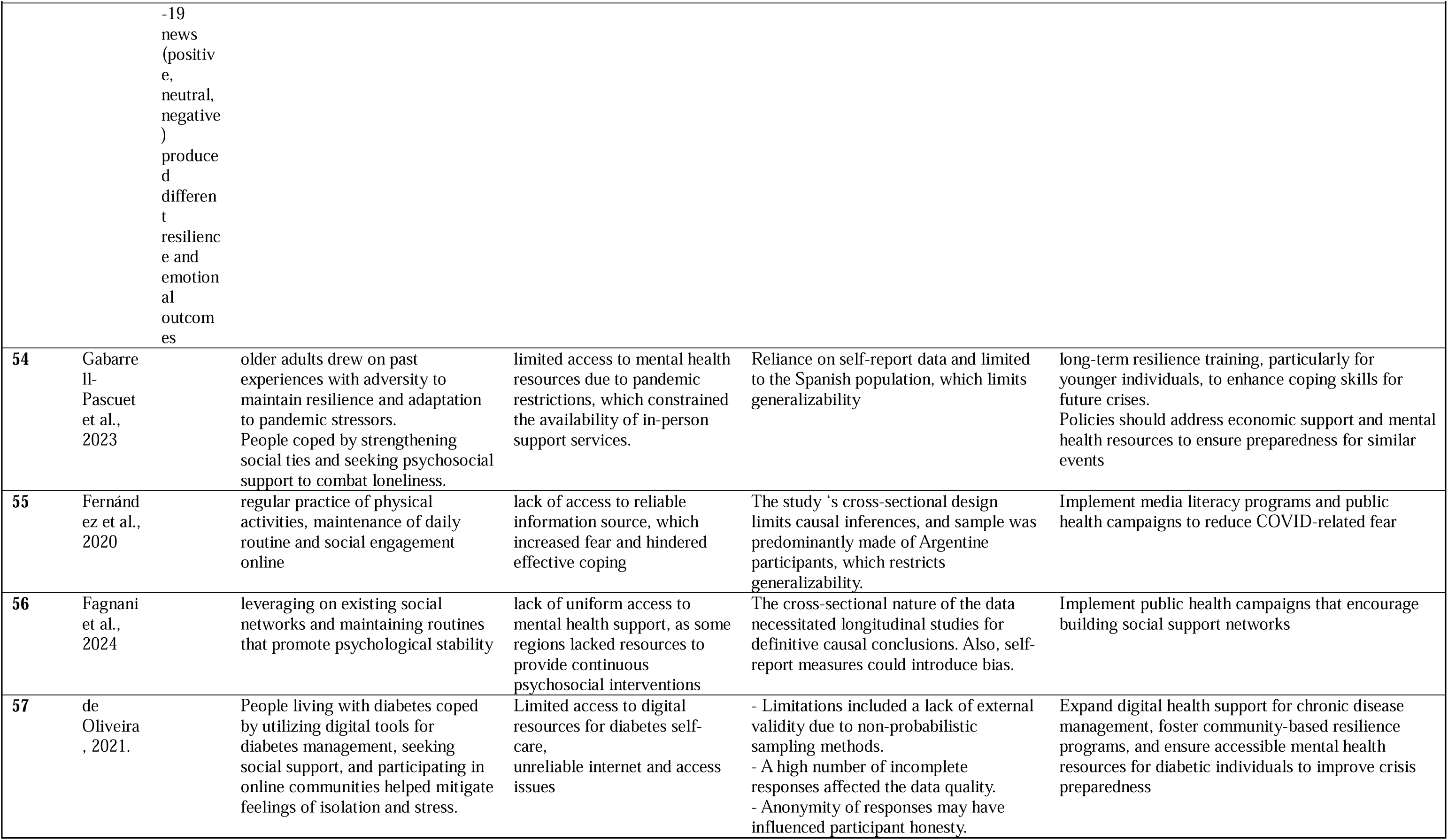
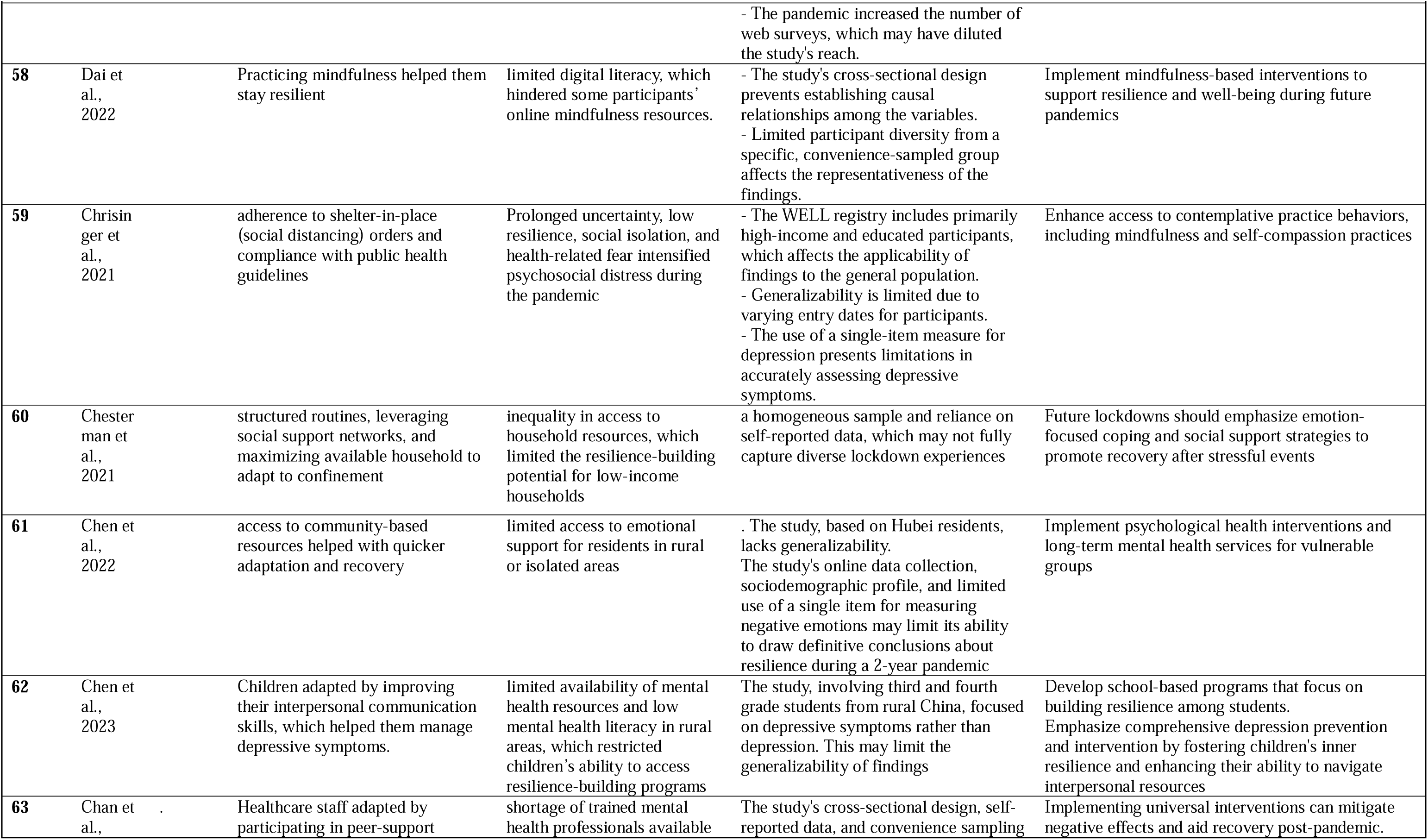
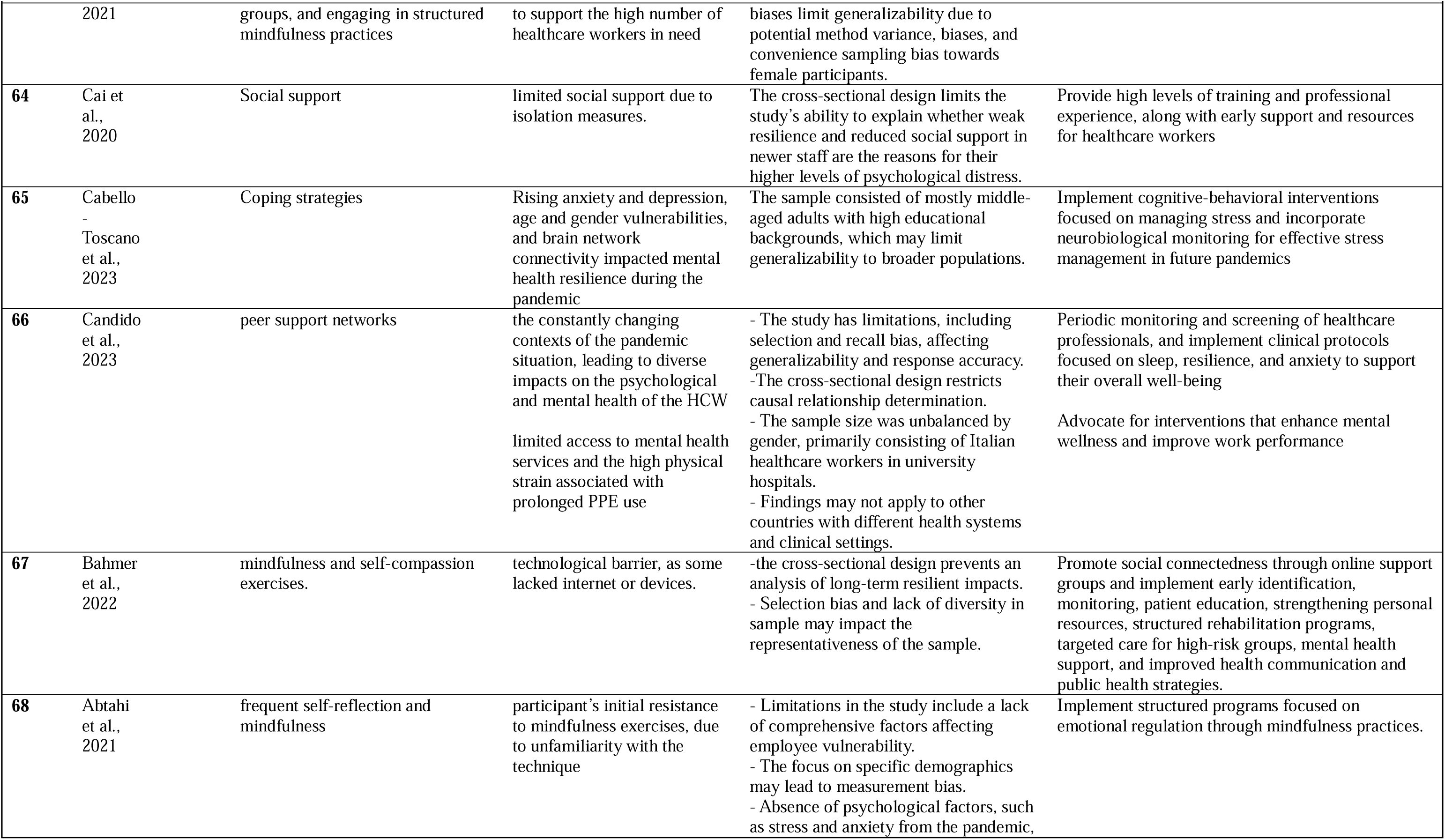
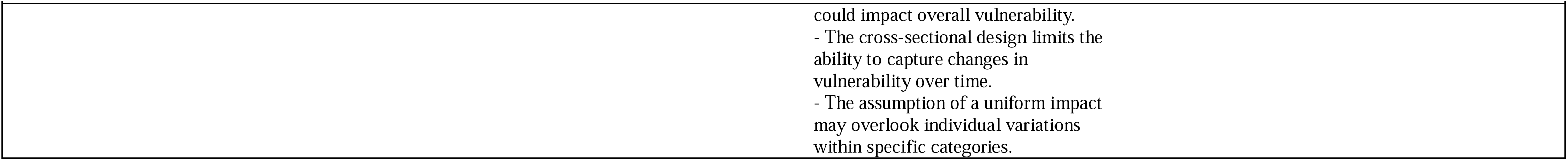
Key lessons from the included studies.

These interventions generally showed promising efficacy in enhancing resilience and mental health outcomes. For example, Mindfulness and emotional intelligence training improved emotional regulation and stress management in adolescents, fostering higher resilience outcomes by equipping them with adaptive coping mechanisms to handle stress adversity^69^. In the experimental group, resilience scores increased over the 6-month period, with significant differences between pre- and post-intervention assessments. Specifically, at T2 (2 months post-intervention), the resilience scores of the experimental group showed a mean increase of 0.64 (SD = 0.25), with t = 24.72, p < 0.001. This positive trend continued at T3 (4 months) and T4 (6 months), with improvements of 0.26 (SD = 0.03) and 0.62 (SD = 0.10), respectively (p < 0.001). The control group showed no significant changes in resilience over the same period ^69^.

The PILOT program for elementary students targeted key behavioral outcomes, including cognitive reappraisal, substance resistance self-efficacy, and psychological flourishing. This intervention contributed to enhanced resilience outcomes while also reducing depressive symptoms and improving other positive behavioral outcomes, such as emotional regulation and tendencies toward internet addiction^67^. Students in the high-exposure group showed higher resilience scores post-intervention compared to those in the low-exposure group. Specifically, the high-exposure group had an adjusted mean resilience score of 23.71 (SD = 7.98), while the low-exposure group had an adjusted mean of 20.25 (SD = 8.71), with a significant difference between the two groups (t = 2.314, p = 0.021) ^67^.

Media exposure was found to influence systemic and behavioral factors associated with resilience. Positive media exposure promoted greater resilience outcomes by increasing positive emotions and reducing negative emotional states, highlighting the role of mass media in fostering resilience at the population level ^59^. Participants exposed to positive news had higher resilience scores (M = 30.61, SD = 4.879) compared to those exposed to negative news (M = 27.39, SD = 6.071). The difference between the positive news and the negative news groups was statistically significant (F = 6.234, p = 0.002), indicating that positive news exposure contributed to higher resilience during the pandemic, whereas negative news had a detrimental effect on resilience^59^

## 4. DISCUSSION

### 4.1. Knowledge gaps

Despite the insightful strategies to promote resilience, the current review has identified several critical knowledge gaps in the fields primarily related to the conceptualization of resilience, study design, measurement tools, sample size, and cultural sensitivity that may impact the robustness and generalizability of the findings.

While resilience was broadly characterized as the ability to positively adapt to stress, adversity, or crises, its conceptualization varied across studies depending on the context in which it was defined. This variability may undermine the generalizability of findings and hinder the development of unified strategies to promote resilience. The quality assessment further emphasizes challenges in the robustness of the included studies, particularly the predominance of cross-sectional designs (n=49) and the limited use of power analysis and sample size justification in both cross-sectional (43/49) and longitudinal studies (10/16). Additionally, a significant number of studies failed to report participation rates, control for confounding variables, or examine varying levels of exposure to resilience factors, further restricting the ability to draw definitive conclusions. These methodological gaps highlight the critical need for standardization in resilience research, particularly in the operationalization of the construct and study design, to improve reliability, cultural sensitivity, and the broader applicability of findings across diverse populations.

The reliance on self-reported measures may have introduced response biases, particularly in studies where resilience and other key variables were evaluated without objective measures or clinical assessment. Furthermore, a variety of measurement tools were used across studies, leading to inconsistencies in how resilience was assessed and making cross-study comparisons challenging.

Sample size and representativeness were additional concerns, as many studies had small or homogeneous samples, often restricted to specific populations, such as HCWs, students, or single geographic regions. This limited the generalizability of findings to broader and more diverse populations. Also, vulnerable populations, such as minority and marginalized groups, were underrepresented, limiting the applicability of findings to these demographics. As these studies were conducted during the pandemic, many of them faced restrictive environments, with data collection often dependent on internet-based surveys. This reliance likely excluded individuals with limited digital access, such as older adults and those in rural areas, introducing selection bias and further limiting generalizability.

The lack of cultural sensitivity in some studies was another notable knowledge gap. Many resilience measurement tools and interventions were not adapted to participants’ cultural or linguistic backgrounds. This lack of cultural adaptation may have impacted the efficacy of interventions and the accuracy of resilience assessments, particularly in non-Western contexts. Finally, the absence of control groups in studies hindered the ability to evaluate intervention efficacy rigorously, leaving potential effects unquantified. These limitations suggest the need for future research with more diverse samples, longitudinal study designs, culturally adapted tools, and control group comparisons to enhance the reliability, applicability, and scope of resilience studies in crisis contexts.

Finally, the number of resilience promotion interventions during and after the pandemic was small, and the three included interventional studies had notable limitations. First, the homogenous nature of the sample populations in these studies limits the generalizability of their findings to broader or more diverse groups. None of the interventions targeted essential workers (e.g., HCWs or teachers) who should be paid large attention in terms of psychosocial well-being during the pandemic. Second, the implementation processes of some interventions varied significantly across populations, even within the same study^67^, potentially affecting the consistency and replicability of the results. Third, the efficacy evaluation of these interventions focused on overall resilience outcomes measured by the CD-RISC. Although utilization of the same measurement enabled cross-study comparison, we may lose information regarding context-specific nuances or variations in resilience development dynamics across diverse conditions. Finally, intervention evaluations in some studies did not collect long-term follow-up data from the participants, leaving questions about their sustained impact unanswered.

### 4.2. Recommendations for future research and intervention

The findings of the 68 included studies inform various strategies to enhance psychological resilience and coping capacities for future crises. Future public health crisis preparedness requires integrative efforts at multiple socioecological levels through prioritizing cognitive and coping interventions, public health policies and interventions, as well as strengthening support systems.

#### 4.2.1. Target essential workers during public health emergencies

Essential workers such as HCWs and teachers endure additional stressors during public health emergencies. Improving resilience among this group not only reduces their emotional strain and burnout, but also significantly contributes to healthcare system resilience and community resilience because health facilities and schools always play a critical role in distributing resources (e.g., face masks, sanitizers, free food, etc.). Effective resilience-building programs are needed to provide training on disaster management, burnout detection, and supportive leadership to mitigate stress and improve resilience^39–42, 44–47, 66^. Workplace support should be integrated into the existing system, such as regularly scheduled debriefing sessions and team huddles^109^, accessing digital teaching resources and a structured learning environment^39,49–51^, and participative decision-making ^49^.

#### 4.2.2. Teach and practice tailored coping strategies to address the challenges and barriers of resilience

Existing literature suggests numerous coping strategies at the individual level but highlights the importance of tailoring the content for vulnerable populations. Cognitive techniques like reframing negative thoughts and self-regulation helped participants manage anxiety and uncertainty, building resilience by improving their capacity to cope with stress ^49, 57, 58, 61–63, 71, 76–79, 83, 84, 87, 89, 93–96, 99, 105^. Problem-focused coping strategies, including increasing knowledge of COVID-19 and adopting protective behaviors such as social distancing, helped individuals feel more in control, enhancing resilience by mitigating health-related anxieties^58, 63, 91, 101^. Interventions that address emotional regulation techniques such as mindfulness, meditation, and self-compassion to prepare individuals for future crises are recommended to equip individuals for future crises ^56, 81, 85, 91, 109^. We also suggest promoting adaptive coping strategies like cognitive reappraisal, humor, and acceptance to support mental health under prolonged stress^50, 71, 72, 78, 79, 83, 96^ and positive lifestyle practices, including regular exercise, healthy sleep routines, and stress management, through public health campaigns^42, 52, 61, 88, 95^.

#### 4.2.3. Promote social connections during crisis and strengthen support networks in the digital age

Social support systems, often maintained through virtual connections, played a significant role in helping individuals adapt and enhancing resilience outcomes. Access to peer support and family engagement also provided emotional and practical assistance, especially for parents and children^40, 41, 44, 48, 57, 61, 64, 65, 72–74, 76,77, 82, 90, 101, 104, 105, 109^. Strong family communication and positive family outlook were associated with reduced depression, stress, and anxiety^97^. Therefore, we encourage interventions that engage family members to enhance adaptability and resilience^61, 76, 84^ through expanding training in communication, emotional support, and conflict resolution to improve relationship dynamics during crises. Community-based intervention efforts and public health campaigns to buffer against stress are also needed to foster social support.

On the other hand, using digital tools like social media and virtual platforms facilitated social interactions and preserved connections, which play a critical role in fostering resilience^39, 86, 92, 104^. Platforms for virtual interactions alleviated isolation. Expanding telehealth services and digital therapeutic resources through online counseling and remote support platforms will increase access to mental health resources^53, 55, 60, 71, 72, 75, 77, 88, 92^, such as telehealth and video games, provided constructive distractions and supported mental health ^86^. Intervention apps can incorporate mindfulness, coping techniques, and emotional regulation activities^50, 56, 83, 96^. To optimize the benefits of digital technologies in resilience promotion, we need to address the digital divide by increasing internet access and offering digital literacy programs, especially for older adults and underserved populations^55, 60, 62^.

#### 4.2.4. Enhance long-term investment in mental health and address structural inequities

A few studies highlight long-term investment in mental health to promote individual and community level resilience. Community and government should play a leadership in promoting capacity building and address structural inequities. Community preparedness and efforts, such as outreach programs and structured support initiatives within schools, colleges, and workplaces, were crucial in enhancing psychological resilience during the COVID-19 pandemic. These crisis adaptation programs can focus on life-skills education, stress management, and coping strategies^48–51, 62, 63, 65, 67, 69^. Supporting community-based mental health programs that incorporate cognitive-behavioral and coping techniques for vulnerable groups can strengthen community-level resilience ^55, 64, 72, 74, 77, 93, 98, 99, 102, 103^. Fostering spiritual or religious coping, such as prayer, can provide comfort and resilience during crises^75, 84^.

Government and institutional policies played a pivotal role in enhancing resilience during the pandemic. For example, vaccination campaigns not only provided physical protection but also significantly reduced anxiety, depression, and stress levels among HCWs, fostering resilience through improved psychological well-being^47^. During the pandemic, government should develop transparent prevention and safety protocols, take timely infection control measures, and disseminate clear, balanced public health messaging to reduce fear and enhance compliance^55, 59, 60, 89, 94, 100^. To improve preparedness for future public health emergencies, governments should prioritize mental health services in terms of early detection, resilience training, and access to psychological services^55, 73, 77, 78, 80^, especially allocating resources to underserved populations such as the unemployed and those with disabilities^58, 68, 74, 76, 90, 92, 93^.

In addition, policies addressing structural disparities, such as equitable access to quality housing and outdoor spaces and providing financial aid and job security for low-income populations, were critical in promoting resilience among vulnerable populations during lockdowns ^87, 90, 101^. Therefore, long-term investments in health equities are recommended with a focus on improving health infrastructure, promoting health education to combat misinformation^58, 68, 74, 76, 90, 92, 93^, addressing housing disparities, and enhancing financial security through inclusive economic policies^87, 90, 101^.

### 4.3. Our study limitations

Although our review provides a comprehensive understanding of resilience, factors associated with resilience, recommendations for future preparedness during pandemics, and knowledge gaps in existing literature, we acknowledge several limitations. The decision to limit our search to only four databases may have restricted the scope of the literature review and potentially excluded relevant studies indexed in other databases. Additionally, the inclusion of only English-language, peer-reviewed studies, along with the exclusion of grey literature, may have led to the omission of valuable insights that could have further enriched the findings. For future literature review, we recommend the expansion of search to include a wider range of databases and sources, including grey literature and non-English publications, to capture a more diverse set of studies. Moreover, incorporating studies from underrepresented geographic regions and targeting more interventional studies could enhance the generalizability and depth of findings on psychological resilience in the context of global public health crises.

## 5. CONCLUSION

This review provides valuable insights into the development of psychological resilience during the COVID-19 pandemic across different levels of the social-ecological model to enhance preparedness for future global crises. Findings demonstrate that psychological resilience was associated with adaptive coping mechanisms, social connections, and access to mental health resources, while barriers such as social isolation and systemic disparities hindered resilience. Importantly, this is the first comprehensive review to consider psychological resilience at different levels globally and across diverse populations. These findings will serve as a valuable resource for future preparedness, offering insights to guide research, policy, and intervention strategies aimed at enhancing resilience during future global crises.

## Data Availability

Since this is a systematic review, the data used in this study were obtained from previously published sources. All data supporting the findings of this review are available within the manuscript and its supplementary materials. Additional details can be provided upon request to the authors.

## Acknowledgment

None.

## Author contributions

**Atena Pasha** (Conceptualization, Investigation, Methodology, Project administration, Formal analysis, Writing – original draft, Writing – review and editing), **Abdul-Hanan Saani Inusah** (Data curation, Formal analysis, Writing – original draft, Writing – review and editing), **Jannatun Nayem** (Investigation, Data curation, Formal analysis), **Xiaoming Li** (Conceptualization, Project administration, Supervision, Funding acquisition, Writing – review and editing), **Shan Qiao** (Conceptualization, Project administration, Supervision, Funding acquisition, Writing – review and editing)

---

**Supplementary Table 1.** Search Strategies for Specific Databases.

| Database | Search String |
| --- | --- |
| **PsycINFO** | ((COVID-19 OR "2019 novel coronavirus" OR "SARS-CoV-2" OR "Coronavirus disease 2019" OR "COVID-19 pandemic" OR "COVID-19 response") AND ("psychological resilience" OR "psychological response" OR resilience OR "psychological resilient response" OR "psychological adaptation")) |
| **PubMed** | (("COVID-19"[MeSH Terms] OR "2019 novel coronavirus" OR "SARS-CoV-2" OR "Coronavirus disease 2019" OR "COVID-19 pandemic" OR "COVID-19 response") AND ("psychological resilience"[MeSH Terms] OR "psychological adaptation" OR resilience OR "psychological response" OR "psychological resilient response")) |
| **ScienceDirect** | TITLE-ABS-KEY(("COVID-19" OR "2019 novel coronavirus" OR "SARS-CoV-2" OR "Coronavirus disease 2019" OR "COVID-19 pandemic" OR "COVID-19 response") AND ("psychological resilience" OR "psychological adaptation" OR resilience OR "psychological response" OR "psychological resilient response")) |
| **Web of Science** | TS=("COVID-19" OR "2019 novel coronavirus" OR "SARS-CoV-2" OR "Coronavirus disease 2019" OR "COVID-19 pandemic" OR "COVID-19 response") AND TS=("psychological resilience" OR "psychological adaptation" OR resilience OR "psychological response" OR "psychological resilient response") |

**Supplementary Table 2a.** Quality Assessment for Cross-sectional Studies.

| Studies | D1 | D2 | D3 | D4 | D5 | D6 | D7 | D8 | D9 | D10 | D11 | D12 | D13 | D14 | Total | Overall Rating |
| --- | --- | --- | --- | --- | --- | --- | --- | --- | --- | --- | --- | --- | --- | --- | --- | --- |
| Zheng et al. (2023) | Yes | Yes | Yes | Yes | No | No | No | Yes | Yes | NA | Yes | NA | NA | Yes | 7 | Good |
| Zheng et al. (2021) | Yes | Yes | No | Yes | No | No | No | Yes | Yes | NA | Yes | NA | NA | No | 6 | Good |
| Zhang et al. (2021) | Yes | Yes | Yes | Yes | No | No | No | Yes | Yes | NA | Yes | NA | NA | Yes | 7 | Good |
| Kocjan et al. (2021) | Yes | Yes | NR | Yes | No | No | No | Yes | Yes | NA | Yes | NA | NA | Yes | 6 | Good |
| Wang et al. (2023) | Yes | Yes | Yes | Yes | No | No | No | Yes | Yes | NA | Yes | NA | NA | Yes | 7 | Good |
| Vijayalakshmi et al. (2023) | Yes | Yes | NR | Yes | No | No | No | No | Yes | NA | Yes | NA | NA | No | 5 | Fair |
| Varma et al. (2021) | Yes | Yes | Yes | Yes | No | No | No | Yes | Yes | NA | Yes | NA | NA | No | 7 | Good |
| Vannini et al. (2021) | Yes | Yes | Yes | Yes | No | No | No | Yes | Yes | NA | Yes | NA | NA | Yes | 7 | Good |
| Üngüren & Kaçmaz (2022) | Yes | Yes | NR | Yes | No | No | No | Yes | Yes | NA | Yes | NA | NA | Yes | 6 | Good |
| Tsibidak (2021) | Yes | Yes | NR | Yes | No | No | No | Yes | Yes | NA | Yes | NA | NA | No | 6 | Good |
| Toyama et al.(2023) | Yes | Yes | Yes | Yes | No | No | No | Yes | Yes | NA | Yes | NA | NA | No | 7 | Good |
| Taylor e. al. (2021) | Yes | Yes | NR | Yes | No | No | No | Yes | Yes | NA | Yes | NA | NA | Yes | 6 | Good |
| Skalski et al. (2021) | Yes | Yes | NR | Yes | No | No | No | Yes | Yes | NA | Yes | NA | NA | No | 6 | Good |
| AlKudsi et al. (2022) | Yes | Yes | Yes | Yes | Yes | No | No | Yes | Yes | NA | Yes | NA | NA | Yes | 8 | Good |
| Rotonda et al. (2021) | Yes | No | No | Yes | No | No | No | Yes | Yes | NA | Yes | NA | NA | Yes | 5 | Fair |
| Reichert et al. (2022) | Yes | Yes | NR | Yes | No | No | No | Yes | Yes | NA | Yes | NA | NA | Yes | 6 | Good |
| Pigati et al. (2022) | Yes | Yes | Yes | Yes | Yes | No | No | Yes | Yes | NA | Yes | NA | NA | Yes | 8 | Good |
| Paredes et al. (2021) | Yes | Yes | No | Yes | No | No | No | Yes | Yes | NA | Yes | NA | NA | No | 6 | Good |
| Padmanabhanunni et al. (2023) | Yes | Yes | NR | Yes | Yes | No | No | Yes | Yes | NA | Yes | NA | NA | No | 7 | Good |
| Ouanes et al. (2021) | Yes | Yes | No | Yes | No | No | No | Yes | Yes | NA | Yes | NA | NA | Yes | 6 | Good |
| Gortari, A. B.O. (2023) | Yes | Yes | NR | Yes | No | No | No | No | Yes | NA | Yes | NA | NA | No | 5 | Fair |
| Meynaar et al. (2021) | Yes | Yes | NR | Yes | No | No | No | Yes | Yes | NA | Yes | NA | NA | No | 6 | Good |
| Meyer et al. (2022) | Yes | Yes | NR | Yes | No | No | No | No | Yes | NA | Yes | NA | NA | Yes | 5 | Fair |
| Martin et al. (2021) | Yes | Yes | Yes | Yes | No | No | No | Yes | Yes | NA | Yes | NA | NA | Yes | 7 | Good |
| Manuti et al. (2022) | Yes | Yes | NR | Yes | No | No | No | Yes | Yes | NA | Yes | NA | NA | No | 6 | Good |
| Lo et al. (2023) | Yes | Yes | Yes | Yes | No | No | No | Yes | Yes | NA | Yes | NA | NA | Yes | 7 | Good |
| Liu et al. (2023) | Yes | Yes | Yes | Yes | No | No | No | Yes | Yes | NA | Yes | NA | NA | No | 7 | Good |
| Lim et al. (2021) | Yes | Yes | Yes | Yes | No | No | No | No | Yes | NA | Yes | NA | NA | No | 6 | Good |
| Lassri (2023) | Yes | Yes | NR | Yes | No | No | No | Yes | Yes | NA | Yes | NA | NA | No | 6 | Good |
| Kural & Kovacs (2021) | Yes | Yes | Yes | Yes | Yes | No | No | Yes | Yes | NA | Yes | NA | NA | Yes | 8 | Good |
| Kuhlman et al. (2021) | Yes | Yes | Yes | Yes | No | No | No | Yes | Yes | NA | Yes | NA | NA | Yes | 7 | Good |
| Kubo et al. (2021) | Yes | Yes | NR | Yes | No | No | No | Yes | Yes | NA | Yes | NA | NA | Yes | 6 | Good |
| Koire et al. (2023) | Yes | Yes | NR | Yes | No | No | No | Yes | Yes | NA | Yes | NA | NA | Yes | 6 | Good |
| Kleimeier et al. (2023) | Yes | Yes | NR | Yes | No | No | No | Yes | Yes | NA | Yes | NA | NA | Yes | 6 | Good |
| Karataş & Tagay (2021) | Yes | Yes | NR | Yes | No | No | No | Yes | Yes | NA | Yes | NA | NA | Yes | 6 | Good |
| Kaim et al. (2021) | Yes | Yes | NR | Yes | Yes | No | No | Yes | Yes | NA | Yes | NA | NA | Yes | 7 | Good |
| Hu et al. (2020) | Yes | Yes | Yes | Yes | No | No | No | Yes | Yes | NA | Yes | NA | NA | No | 7 | Good |
| Hébert et al. (2022) | Yes | Yes | NR | Yes | No | No | No | No | Yes | NA | Yes | NA | NA | Yes | 5 | Fair |
| Han et al. (2022) | Yes | Yes | NR | Yes | No | No | No | No | Yes | NA | Yes | NA | NA | No | 5 | Fair |
| Haldorai et al. (2023) | Yes | Yes | NR | Yes | Yes | No | No | Yes | Yes | NA | Yes | NA | NA | Yes | 7 | Good |
| Gizdic et al. (2023) | Yes | Yes | NR | Yes | No | No | No | Yes | Yes | NA | Yes | NA | NA | Yes | 6 | Good |
| Fernández et al. (2020) | Yes | Yes | NR | Yes | No | No | No | Yes | Yes | NA | Yes | NA | NA | Yes | 6 | Good |
| de Oliveira et al. (2021) | Yes | Yes | Yes | Yes | No | No | No | No | Yes | NA | Yes | NA | NA | No | 6 | Good |
| Dai et al. (2022) | Yes | Yes | No | Yes | No | No | No | Yes | Yes | NA | Yes | NA | NA | Yes | 6 | Good |
| Chesterman et al. (2021) | Yes | Yes | NR | Yes | No | No | No | No | Yes | NA | Yes | NA | NA | Yes | 5 | Fair |
| Chan et al. (2021) | Yes | Yes | Yes | Yes | No | No | No | Yes | Yes | NA | Yes | NA | NA | Yes | 7 | Good |
| Cai et al. (2020) | Yes | Yes | NR | Yes | No | No | No | Yes | Yes | NA | Yes | NA | NA | Yes | 6 | Good |
| Candido et al. (2023) | Yes | Yes | CD | Yes | No | No | No | No | Yes | NA | Yes | NA | NA | Yes | 5 | Fair |
| Abtahi et al. (2021) | Yes | Yes | No | Yes | No | No | No | Yes | Yes | NA | Yes | NA | NA | No | 6 | Good |

**Supplementary Table 2b.** Quality Assessments of Longitudinal Studies.

| Studies | D1 | D2 | D3 | D4 | D5 | D6 | D7 | D8 | D9 | D10 | D11 | D12 | D13 | D14 | Total | Overall Rating |
| --- | --- | --- | --- | --- | --- | --- | --- | --- | --- | --- | --- | --- | --- | --- | --- | --- |
| Sun et al. (2021) | Yes | Yes | Yes | Yes | Yes | NA | Yes | NR | No | Yes | No | NR | Yes | Yes | 8 | Fair |
| Spada et al. (2022) | Yes | Yes | Yes | Yes | Yes | Yes | Yes | NR | Yes | Yes | No | NR | CD | Yes | 9 | Good |
| Rossi et al. (2023) | Yes | Yes | Yes | Yes | Yes | NA | Yes | NR | No | NA | No | NR | No | Yes | 6 | Fair |
| Na et al. (2022) | Yes | Yes | Yes | Yes | Yes | NA | Yes | NR | Yes | Yes | Yes | NA | Yes | Yes | 10 | Good |
| Na & Yang, (2022) | Yes | Yes | NR | Yes | No | Yes | Yes | Yes | Yes | Yes | Yes | Yes | NR | No | 10 | Good |
| Letcher et al. (2023) | Yes | Yes | NR | Yes | No | Yes | Yes | Yes | Yes | Yes | Yes | NR | Yes | Yes | 10 | Good |
| Kimhi et al. (2021) | Yes | Yes | NR | Yes | No | Yes | Yes | NR | Yes | Yes | Yes | NR | No | Yes | 8 | Fair |
| Giusti et al. (2023) | Yes | Yes | Yes | Yes | No | Yes | Yes | Yes | Yes | Yes | Yes | NR | Yes | Yes | 11 | Good |
| G-Pascuet et al. (2023) | Yes | Yes | Yes | Yes | Yes | Yes | Yes | Yes | Yes | Yes | Yes | NR | Yes | Yes | 12 | Good |
| Fagnani et al. (2024) | Yes | Yes | No | Yes | No | Yes | Yes | Yes | Yes | Yes | Yes | NR | No | Yes | 9 | Good |
| Chrising er et al. (2021) | Yes | Yes | NR | Yes | No | Yes | Yes | CD | Yes | Yes | Yes | NR | Yes | No | 9 | Good |
| Chen et al. (2022) | Yes | Yes | NR | Yes | No | Yes | Yes | Yes | Yes | Yes | Yes | NR | NA | No | 9 | Good |
| Chen et al. (2023) | Yes | Yes | Yes | Yes | No | Yes | Yes | Yes | Yes | Yes | Yes | NR | Yes | Yes | 11 | Good |
| CabelloToscano et al. (2023) | Yes | Yes | NR | Yes | No | Yes | Yes | Yes | Yes | Yes | Yes | NR | NR | Yes | 9 | Good |
| Bahmer et al. (2022) | Yes | Yes | NR | Yes | Yes | Yes | Yes | Yes | Yes | Yes | Yes | NR | NR | Yes | 10 | Good |
| Urmanche et al. 2023 | Yes | Yes | NR | Yes | No | Yes | Yes | Yes | Yes | Yes | Yes | NR | No | No | 9 | Good |

**Supplementary Table 2c.** Quality Assessment of Experimental Studies.

| **Studies** | **D1** | **D2** | **D3** | **D4** | **D5** | **D6** | **D7** | **D8** | **D9** | **D10** | **D11** | **D12** | **D13** | **D14** | **Total** | **Overall Rating** |
| --- | --- | --- | --- | --- | --- | --- | --- | --- | --- | --- | --- | --- | --- | --- | --- | --- |
| Yuan, (2021) | Yes | Yes | CD | NR | NR | Yes | Yes | Yes | Yes | Yes | Yes | No | Yes | No | 9 | Good |
| Giri et al. (2021) | Yes | Yes | NR | NR | NR | Yes | Yes | NR | Yes | Yes | Yes | Yes | Yes | No | 9 | Good |
| Wu et al. (2023) | Yes | No | Yes | NR | NR | Yes | Yes | Yes | Yes | Yes | No | NA | - | - | 7 | Good |

